# Investigating Dopaminergic Abnormalities in Psychosis with Normative Modelling and Multisite Molecular Neuroimaging

**DOI:** 10.1101/2023.11.27.23299051

**Authors:** A. Giacomel, D. Martins, G. Nordio, R. Easmin, O. Howes, Pierluigi Selvaggi, S.C.R. Williams, F. Turkheimer, M. De Groot, O. Dipasquale, M. Veronese, the FDOPA PET imaging working group

## Abstract

Molecular neuroimaging techniques, like PET and SPECT, offer invaluable insights into the brain’s in-vivo biology and its dysfunction in neuropsychiatric patients. However, the transition of molecular neuroimaging into diagnostics and precision medicine has been limited to a few clinical applications, hindered by issues like practical feasibility and high costs. In this study, we explore the use of normative modelling (NM) for molecular neuroimaging to identify individual patient deviations from a reference cohort of subjects. NM potentially addresses challenges such as small sample sizes and diverse acquisition protocols that are typical of molecular neuroimaging studies. We applied NM to two PET radiotracers targeting the dopaminergic system ([^11^C]-(+)-PHNO and [^18^F]FDOPA) to create a normative model to reference groups of controls. The models were subsequently utilized on various independent cohorts of patients experiencing psychosis. These cohorts were characterized by differing disease stages, treatment responses, and the presence or absence of matched controls. Our results showed that patients exhibited a higher degree of extreme deviations (∼3-fold increase) than controls, although this pattern was heterogeneous, with minimal overlap in extreme deviations topology (max 20%). We also confirmed the value of striatal [^18^F]FDOPA signal to predict treatment response (striatal AUC ROC: 0.77-0.83). Methodologically, we highlighted the importance of data harmonization before data aggregation. In conclusion, normative modelling can be effectively applied to molecular neuroimaging after proper harmonization, enabling insights into disease mechanisms and advancing precision medicine. The method is valuable in understanding the heterogeneity of patient populations and can contribute to maximising cost efficiency in studies aimed at comparing cases and controls.

## Introduction

Psychosis is a debilitating mental health condition characterised by a loss of contact with reality, abnormal thoughts, perceptions, and behaviour. While psychosis is a hallmark of schizophrenia, psychotic symptoms can also be present in other psychiatric disorders including bipolar disorder, and severe depression. Psychosis has been the subject of extensive neuroimaging research aimed at unravelling its underlying neurobiological mechanisms. Consistent findings from structural magnetic resonance imaging (MRI) studies have systematically shown brain structure abnormalities, such as reduced grey matter volume and compromised white matter integrity^1^. Similarly, functional MRI (fMRI) studies have identified abnormal connectivity and atypical brain activation patterns during cognitive tasks, suggesting dysfunction in cortical and subcortical regions associated with attention, memory, and emotion regulation^2^. Molecular neuroimaging, such as Positron Emission Tomography (PET), has also provided valuable insights into the molecular alterations associated with psychosis, highlighting the involvement of multiple neuroreceptor systems, metabolism^3^ and neuroinflammation^4^.

Abnormalities in dopamine neurotransmission have emerged as a consistent finding in psychosis^5^. The dopamine hypothesis posits dysregulation within the dopaminergic system as a key factor in the development of psychosis^6^. PET studies have consistently shown elevated dopamine synthesis capacity in specific brain regions, particularly the striatum, in individuals with psychosis^7^. Specifically, this hyperactivity of the striatal dopaminergic system is believed to contribute to positive symptoms such as hallucinations and delusions^8–10^. The glutamate hypothesis has also gained significant attention in psychosis research. This hypothesis suggests dysfunction in the glutamatergic system, particularly N-methyl-D-aspartate (NMDA) receptor hypofunction, as a contributing factor for psychosis^11,12^. [^18^F]FDOPA PET studies have also provided evidence of altered dopamine-glutamate interactions, highlighting the intricate relationship between these neurotransmitter systems^5^.

While offering unique insights into the brain mechanisms underlying psychosis, most of the neuroimaging literature has primarily focused on attempting to identify single unifying pathophysiological processes shared across patients. This has been achieved using ‘standard’ cross-sectional statistics based on group averages, often treating individual differences merely as noise. However, there is a growing awareness that these conventional approaches fall short of fully capturing the multifaceted characteristics of complex mental health disorders like psychosis. To develop imaging-based biomarkers with true clinical utility, interindividual variability cannot be simply dismissed as noise or assumed to be part of measurement variability. This awareness has been fostered, in part, by the parallel tendency of collecting large archives of neuroimaging data, which has triggered a gradual shift in neuroimaging towards employing advanced analytical modelling methods to model subjects’ characteristics^13^. By providing statistical inferences at the individual level with respect to an expected pattern, these methods offer the opportunity to parse heterogeneity across cohorts and identify the unique characteristics of each individual. Furthermore, by focusing on personalized perspective and acknowledging the potential to serve as fundamental keys in the creation of neuroimaging biomarkers for furthering understanding of the neurobiological basis of psychiatric disorders and potentially predicting treatment outcomes^14,15^.

One of these advanced modelling methods that has gained widespread traction in neuroimaging research is normative modelling (NM). This statistical framework is based on the concept of paediatric growth charts^16^, which utilise a series of percentile curves to illustrate the normal distribution of children’s body measurements, such as weight, height, and head circumference, as a function of their age. When applied to neuroimaging data^14,17,18^, this approach enables the identification of a relationship between quantitative neuroimaging biomarkers, such as regional brain volume or thickness measured with MRI, and relevant factors like specific clinical, demographic, or behavioural measures of interest^17^.

The rationale for employing normative modelling in neuroimaging is twofold. Firstly, this approach allows us to use neuroimaging data from healthy individuals to establish the normal range for a specific brain characteristic. By describing a portion of the between-subject variability of the given neuroimaging measure through demographic factors (e.g., age, sex^19,20^, BMI) or abilities (e.g., IQ^21^), normative modelling defines what can be considered typical or within the range of expected variation. Secondly, it allows for the quantification of regional deviations from normality at the individual level. This aspect aims to identify disease-specific patterns of alterations and dissect disease heterogeneity across patients. By comparing an individual’s neuroimaging data to the established normal range, it is possible to pinpoint aberrations that may indicate the presence of certain conditions and contribute significantly to the understanding and characterization of brain disorders^14,22^.

Up to now, normative modelling has found extensive application in the identification of consistent disease-specific patterns of brain structural alterations, as demonstrated in schizophrenia^19,20,23^, bipolar disorder^19,20^, ADHD^24^, and Parkinson’s disease^25^, using structural MRI data (i.e., T1-weighted MRI^19^ and diffusion-weighted imaging^26^), However, this methodology has yet to be applied to the molecular underpinnings of human brain function. The primary challenge in applying normative modelling to PET/SPECT brain imaging lies in the necessity of pooling large datasets to establish reliable parameters for the normative models. To estimate normative models of MRI-based brain measures, hundreds^19,24,27^ or even thousands^28^ of scans are typically included in the reference cohort. However, these numbers cannot be feasibly achieved in single molecular neuroimaging studies due to the substantial costs of PET and SPECT scans (up to 10 times higher than MRI scans) and ethical issues related to the use of radioactive tracers for research purposes. Nevertheless, recent developments in the molecular neuroimaging community, including a greater willingness to share data^29,30^ and the establishment of international consortia (e.g., ENIGMA^31^), along with the development of effective harmonisation techniques for neuroimaging data^32–35^, have paved the way for the use of normative modelling in molecular neuroimaging.

In this work, our primary objective is to demonstrate the feasibility of employing normative modelling to molecular neuroimaging in the context of psychosis studies. Despite the challenges posed by small sample sizes and diverse experimental designs, we hypothesise that normative modelling can be effectively applied to this type of data, providing valuable insights into molecular brain functions in healthy individuals and those experiencing psychosis. Furthermore, we hypothesise that normative modelling can serve as a powerful tool to identify both the magnitude and the spatial distribution of molecular alterations at the individual patient level by 1) extending analysis of dopamine alterations in the striatum to the whole brain, 2) identifying common patterns of dopamine dysfunction across multiple and independent datasets of patients with psychosis, and 3) linking inter-individual deviation from normalities to clinical symptoms and response to treatment.

This study is hence organised into two main parts. In the first section, we used two datasets^36,37^ of [^11^C]-(+)-PHNO PET imaging data measuring D_2/3_ dopamine receptor density, acquired with two scanners, to compare the effects of two image harmonisation methods on the distribution of the deviation scores of the NM and evaluated which one is the most effective at reducing the scanner effects. We then applied the so-identified optimal harmonisation method to studies of dopamine synthesis capacity using [^18^F]FDOPA PET imaging in healthy controls (HC), acquired with five different scanners, and assessed if the [^11^C]-(+)-PHNO results were generalizable to a different radiotracer and a greater number of scanners.

In the second section, we used the estimated [^18^F]FDOPA PET model to calculate the deviations from normality in four datasets^38–47^ acquired in patients with psychosis, one of which did not include any matched HC. Here, we investigated the presence of shared spatial patterns of deviation in psychosis, but also differences between patients with first-episode psychosis and chronic schizophrenia. In addition, we tested the feasibility of using independent datasets of patients with no matched HCs when a big enough reference cohort is already available. Here, we assessed the replicability of the findings from the previous comparisons and investigated the clinical value of extreme deviations by looking at relationships between patient-specific extreme deviations from normality and clinical symptoms. Lastly, since [^18^F]FDOPA PET imaging has been recently proposed as a potential biomarker for treatment stratification in psychosis^38^, we built a classifier to assess the value of [^18^F]FDOPA PET NM for predicting treatment response, and compared its performances to the reference standard analytics, to evaluate if these would be comparable or outperforming.

## Methods

#### Box 1: Normative Modelling Theory

The general framework to derive normative models from neuroimaging data is explained in detail elsewhere^14,17,22^. In brief, it comprises several steps (Figure B1). Firstly, a reference cohort of healthy controls (HC) is chosen. Next, a specific brain measure, such as a given summary measure at the whole-brain level, region-of-interest (ROI)-based derivatives or voxel-wise brain data from structural MRI scans (e.g., volume and cortical thickness), is selected. Additionally, a set of variables (i.e., predictors) is chosen to explain the brain measure. Statistical models are then constructed for each summary measure, establishing a connection between the neuroimaging data and the selected predictors. The primary outcome of the NM is a measure of deviation, typically expressed as a z-score. This z-score indicates the extent to which the specific brain measure deviates from the normative distribution, providing valuable information about its deviation from the reference group. Following the model’s estimation, its ability to link individual covariates to neuroimaging data needs to be further evaluated both in-sample (e.g., using k-fold cross-validation) and out-of-sample (e.g., using an independent cohort of HC)^22^. Once the model is validated, it can be used to estimate the deviations of a target cohort (generally corresponding to patients), which can then be analysed by investigating the magnitude and spatial pattern of the so-called “extreme deviations”. These extreme values represent brain areas deviating more than two standard deviations from the reference mean.

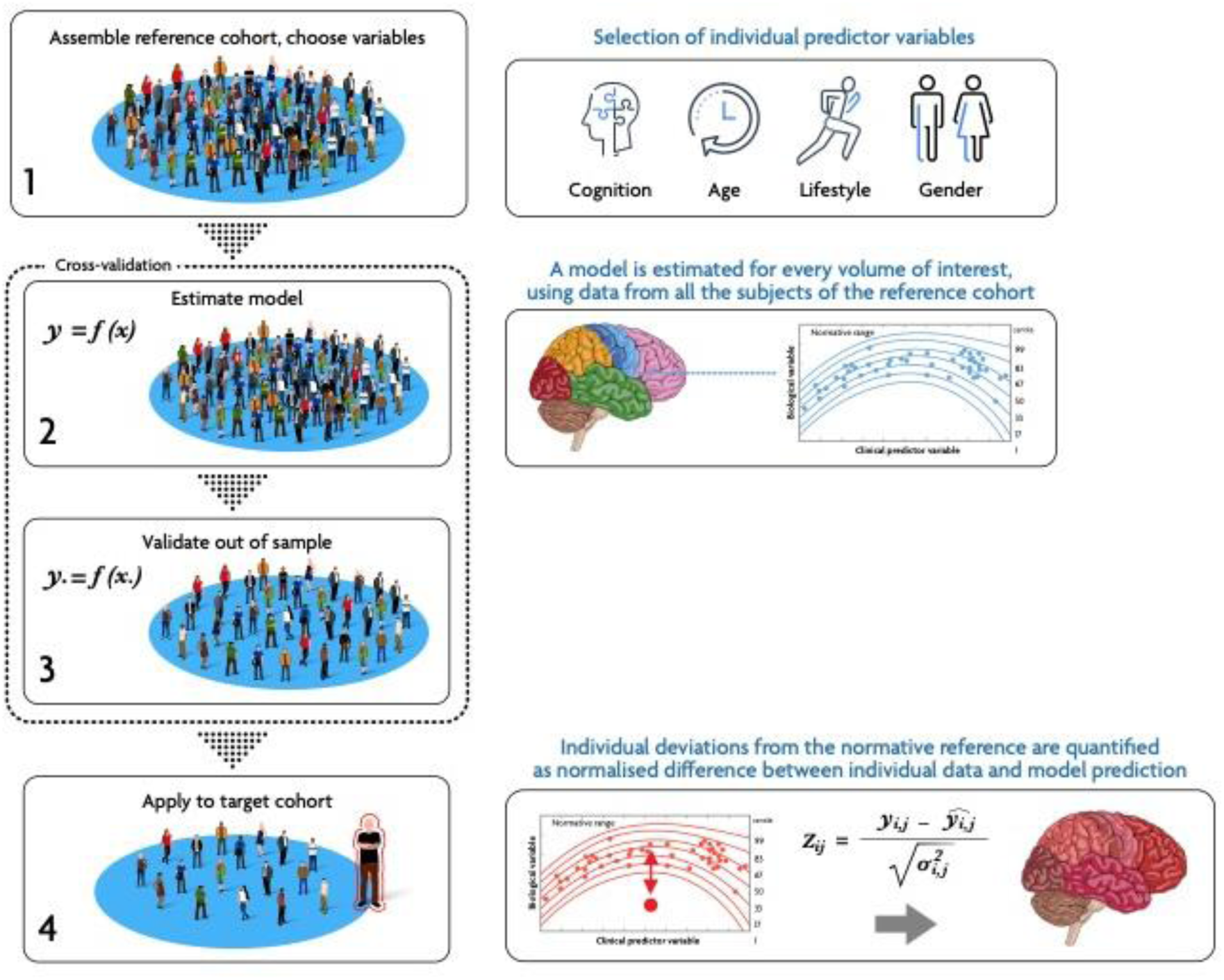
Steps of normative modelling. Main steps for the generation and application of normative modelling to neuroimaging data. In brief, the main steps include (1) the assembly of a reference cohort (usually HC) and the selection of covariates of interest (e.g., demographics or cognitive). (2) Model estimation, at desired level of granularity (i.e., voxel- or ROI-level). (3) Model validation, if possible, out-of-sample, or using cross-validation. (4) Application to target cohort, usually patients, and estimation of deviation scores (Z-scores).

### Study datasets

#### [^11^C]-(+)-PHNO Datasets

Data from 77 HC (Table 1) from two previous studies^36,37^, acquired with [^11^C]-(+)-PHNO, were used to assess and select the best image harmonisation method. The scans were acquired with two different scanners (PET/CT, Siemens Hi-Rez Biograph 6, N=54; and PET/MR, Siemens Biograph mMR, N=23) in the same imaging site (Invicro, London).

**Table 1.**
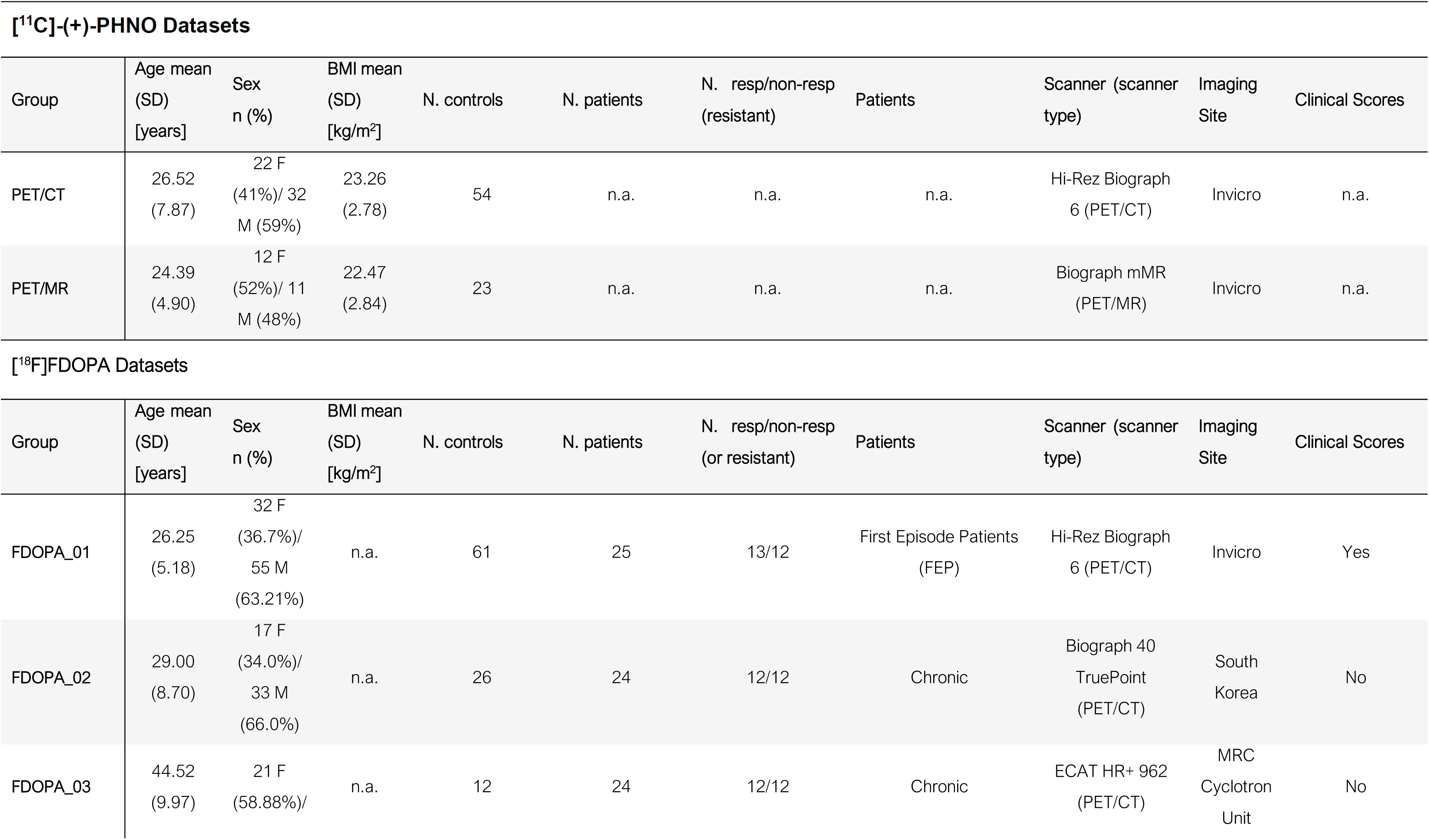
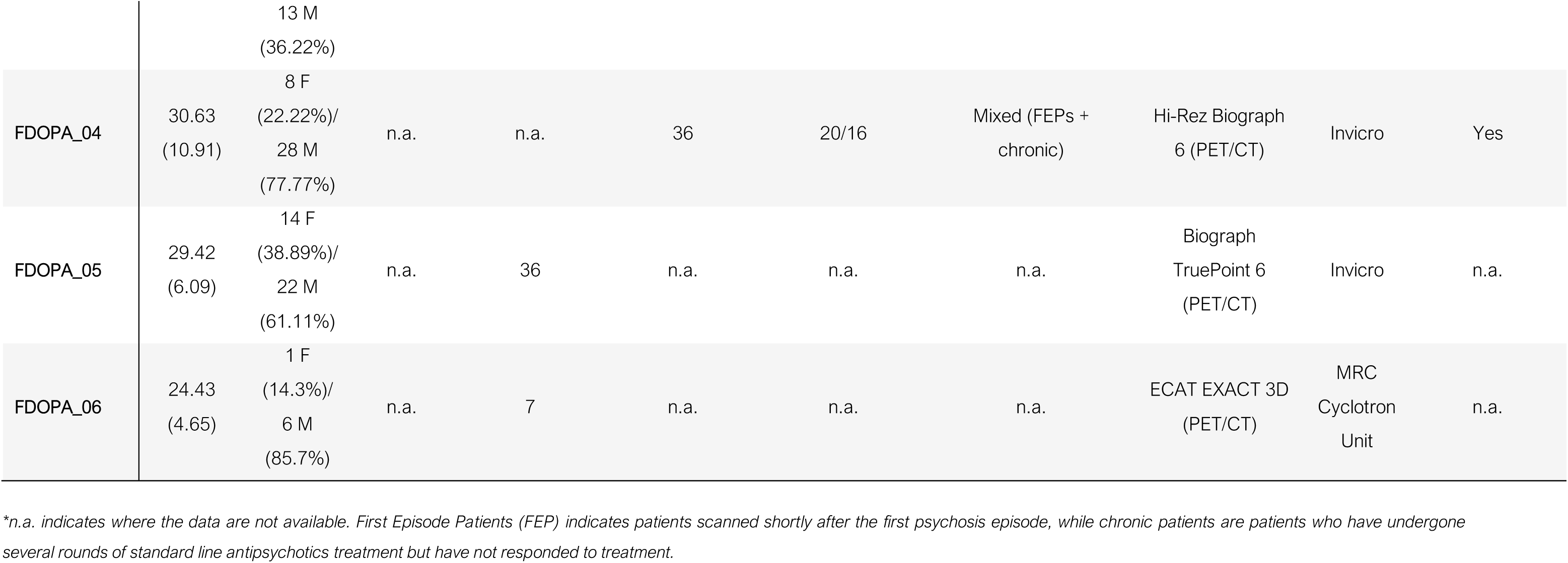
Study datasets.

Experimental designs and imaging protocols were consistent between the two studies. Further details on data acquisition, image processing, and data analysis are reported in the original publications^36,37^. In short, dopamine D2/3 receptor density was measured using non-displaceable binding potential (BP_ND_^48^) as the parameter of interest. For both scanners, parametric BP_ND_ images were obtained using the same MATLAB-based pipeline with a simplified reference tissue model with cerebellar grey matter used as the reference region. Final maps were all normalised to MNI152 standard space before image harmonisation and NM.

#### [^18^F]FDOPA Datasets

6-[^18^F]-fluoro-L-DOPA (FDOPA) data from 142 HC and 109 patients with psychosis from previous studies^38–47^ were used for estimating the NM of dopamine synthesis capacity and exploration of its alterations in psychosis (Table 1). The datasets consisted of two datasets of HC only (FDOPA_HC01 and FDOPA_HC02), three case-control datasets (FDOPA_01, FDOPA_02, and FDOPA_03) and one dataset of patients only (FDOPA_04). Data were acquired with five different scanners (Siemens Hi-Rez Biograph 6, Siemens Biograph 40 TruePoint, Siemens TruePoint 6, ECAT HR+ 962, and ECAT EXACT 3D) in three different imaging sites (MRC Cyclotron Unit, London; Invicro, London and Bundang Hospital, South Korea). The acquisition protocol was consistent across sites. All FDOPA PET imaging sessions were acquired with a continuous dynamic acquisition (no blood sampling), with scanning beginning with the tracer injection and lasting for ∼90 minutes. All participants received carbidopa (150 mg) and entacapone (400 mg) orally 1 hour before imaging to increase the brain tracer uptake and reduce the peripheral formation of radiolabelled metabolite, respectively. The FDOPA tracer (injected dose ranging from 86.4 to 414.4 MBq,) was administered by intravenous bolus injection after the acquisition of a brain CT or MRI for attenuation correction, depending on the scanner availability at each imaging site. PET data reconstruction varied across imaging sites and scanner types, but all included correction for random noise, scatter, and tissue attenuation. Dopamine synthesis capacity indicated by the parameter K_i_^cer^ (sometimes indicated as K_i_, min^-1^) was quantified with the Gjedde-Patlak graphical method using the cerebellum as the reference region. Data were normalised to MNI152 standard space using the same in-house pipeline^49^, removing data with poor quality and excess of motion.

Patients consisted of individuals experiencing either a first episode of psychosis (FEP) or diagnosed with a chronic psychotic disorder (Table 1). In brief, FDOPA_01 patients were recruited if they met structured clinical interview for DSM-IV axis I disorder criteria for schizophrenia (mean Total PANSS [SD]: 72.94 [16.46]), had at least one current or previous psychotic episode (duration of illness, median [IQR]: 24 [24] months), and were antipsychotic medication naïve (no current or previous treatment) or free (not taking medication at scanning time with at least 6 months washout for oral medication or 6 months for depot medication)^39^. FDOPA_02 patients were composed of patients with chronic illness (duration of illness, mean [SD]: 128 [112.24] months) and were recruited if they met the following criteria: they met the DSM-IV criteria for schizophrenia, had a total score ≤ 80 in the PANSS Total scale (mean Total PANSS [SD]: 50.0 [9.64]), had received first-line antipsychotic drugs (including risperidone, olanzapine, and paliperidone) or clozapine for at least 12 weeks^43^. Patients in the FDOPA_03 cohort were also patients with chronic illness who met the DSM-IV criteria for schizophrenia (duration of illness, mean [SD]: 193.8 [367.08] months), received at least two sequential antipsychotic trials (of at least 4 weeks duration), had a Total PANSS score ≥ 75, and were not taking clozapine at time of scanning^8^. Lastly, FDOPA_04 patients were included if they met the DSM-V criteria for schizophrenia or schizophreniform disorders, and were not medicated with clozapine in the previous 3 months to scanning^44^. Full information on the recruitment criteria and clinical characteristics of the patient population is reported in the original papers^38–47^.

For a subset of patients of the cohorts FDOPA_01 and FDOPA_04, symptom severity measures (i.e., Positive and Negative Symptoms Scale – PANSS^50^) were also available.

### Data harmonisation strategies

To harmonise the [^11^C]-(+)-PHNO scans we employed two methods widely used in the literature: Gaussian kernel smoothing^34^ and Combat harmonisation^32,33^.

<u>Gaussian kernel smoothing</u> (hereafter referred to as “smoothing”): This method involves applying a 3D convolution filter with a Full Width Half Maximum (FWMH) of 2.35σ, where σ represents the kernel size. For this step, we used FSL maths^51–53^ setting a kernel size of 3mm, determined through iterative analysis to achieve the best match between the reference and the target data to be harmonised (i.e., PET/CT and PET/MR, respectively).

#### Combat harmonisation

Combat is a Bayesian harmonisation method derived from the field of genomics^54^, which assumes site-specific scale- and shift-factors to be estimated between different batches (e.g., scanners or sites). Unlike smoothing, Combat allows to estimate and preserve the effect of certain covariates defined a priori on the data variability. In our specific case, we employed the NeuroCombat python library (v.0.2.10+)^32,33^ and preselected the same covariates subsequently used for NM (i.e., age, sex for both tracers and BMI for [^11^C]-(+)-PHNO).

To assess the best harmonisation method, we evaluated the effects of harmonisation on the NM results, particularly focusing on the Z-score distributions and the variance explained by the model (see section ‘Comparison of the harmonisation methods’ below).

### NM implementation

After data harmonisation, Bayesian Linear Regression (BLR) was used to estimate voxel-wise NMs (i.e., one model per voxel) of dopamine receptor density and dopamine synthesis capacity from [^11^C]-(+)-PHNO and [^18^F]FDOPA data of the HC samples. BLR is a probabilistic approach to linearly model the relationship between a dependent variable and a set of independent variables. In this case, the selected independent variables were sex, age, and BMI for [^11^C]-(+)-PHNO, and age and sex for [^18^F]FDOPA. Of note, these covariates were used in previous in-house analyses after assessing them to be significant for the specific tracers^49,55–57^.

NMs were estimated using the PCNToolkit (v.1.20.0) Python library, employing default non-informative priors for the parameters and the Powell optimizer for faster convergence of model fitting^14^. Consistently with the PET signal distribution, NM was restricted to the grey matter voxels by thresholding and binarizing a standard probabilistic map of the grey matter at 30% probability. Thresholding for GM is not typically applied for FDOPA brain PET imaging^49^, but this threshold was used as a trade-off between brain coverage and the computational cost of NM quantification. As a result, only voxels with at least 30% probability of being grey matter were included in the mask, yielding a total of 199,715 voxels (and hence models) for each tracer. Importantly, for the [^11^C]-(+)-PHNO data we estimated three different NMs, i.e., one for the unharmonized data and one for each harmonisation strategy (smoothing and Combat) to identify the strategy that best minimises residual scanner-related differences for PET data. Based on the outcome of this analysis of performance, we then used the identified method to harmonise the [^18^F]FDOPA datasets before the NM step.

The estimated NMs were then used to estimate the voxel-wise maps of deviation from normality for each individual. These maps, expressed in terms of Z-scores, measure the distance of a given data point in relation to the average and standard deviation of the posterior probability, weighted for the subject-specific covariates. These values were estimated using k-fold cross-validation (k=5) for the HC, or the full [^18^F]FDOPA NM for patients with psychosis. Furthermore, we identified voxels with an extreme deviation from normality (i.e., voxels with PET signal intensity significantly different from the reference distribution). Such extreme deviations were defined using a thresholding approach: Z>2 for *extreme positive deviations*, Z<-2 for *extreme negative deviations*, and |Z|>2 for *total extreme deviations*. As the deviation scores follow a normal distribution, it is essential to note that there is an expected percentage of extremely deviating voxels. This consists of 2.5% residual density on each tail of the distribution (i.e., Z>2 or Z<2), totalling 5% for the absolute value (i.e., |Z|>2). Conversely, we anticipate a higher percentage of extremely deviating voxels, either positive or negative, in subjects deviating from normality.

### Identification of the optimal harmonisation strategy based on [11C]- (+)-PHNO data

For each harmonisation strategy (no harmonisation, harmonisation with smoothing and harmonisation with combat), we compared the average and standard deviation of the individual Z-score distributions between the two scanners (i.e., PET/CT and PET/MR) to identify any residual differences between datasets. These comparisons were performed using Welch’s two-sample t-tests, implemented with the rstatix package^58^ in R (version 4.2.1). Furthermore, using the brain parcellation defined by the Hammersmith atlas^59^, for each harmonisation strategy we estimated the regional percentage of extreme positive and negative deviations across subjects of each dataset (i.e., PET/CT and PET/MR data). We then calculated the regional differences between the two datasets and employed Wilcoxon’s one-sample rank-tests implemented with the rstatix package^58^ in R 4.2.1 to identify any significant scanner-related differences.

### Validation of the optimal harmonisation strategy on [^18^F]FDOPA data

We validated the harmonization method identified as optimal for the [^11^C]-(+)-PHNO data on the [^18^F]FDOPA data, to see if it would yield consistent results in terms of removing any scanner-related differences across datasets.

First, we harmonised the [^18^F]FDOPA HC data with the optimal harmonisation strategy identified using the [11C]-(+)-PHNO data. Then, in order to include in the analysis only those voxels for which the NMs were able to describe the between-subject variability based on the variables used to estimate the model (i.e., age and sex for the [^18^F]FDOPA data), we discarded the grey matter voxels where NMs were not able to converge to a solution, i.e., those reporting an explained variance (EXPV) lower than 0. Furthermore, since a positive yet very small EXPV does not necessarily mean that the model is able to meaningfully describe the data, we decided to restrict the subsequent analyses to voxels with EXPV > 3% (i.e., voxels where NM reached statistical significance of p_uncorr_ <0.05). For completeness, we repeated the analyses on the data including voxels with EXPV > 0, and using a conservative threshold of EXPV ≥ 10%, corresponding to the optimal point from the L-curve distribution of EXPV.

Finally, to validate the optimal harmonisation strategy on [^18^F]FDOPA HC data, we calculated the mean and standard deviation of the individual distributions of HC Z-score deviations and compared them by means of one-way ANOVAs using the rstatix^58^ library in R 4.2.1.

### NM of [^18^F]FDOPA PET in psychosis

After calculating the z-score deviations of the three clinical cohorts using the NMs estimated from the HC data, we grouped the Z-score distributions for HC and patients separately and compared them. We also calculated the Risk-Ratios (RR)^60^ by counting the number of voxels whose Z-scores showed extreme deviations from normality, using the epitools library in R 4.2.1. In this case, the RR indicates the likelihood of a new individual of belonging to the clinical cohort as compared to the group of HCs.

We then tested the hypothesis that patients would show a greater overlap of extreme deviations than HC, indicating spatial consistency in the manifestation of the disorder, by investigating the spatial patterns of extreme deviations in HC and patients. This was done by counting, for each voxel and group, the number of subjects showing an extreme deviation in that voxel. This was assessed both on the positive and negative extreme deviations separately and on the total extreme deviations.

We then analyzed the differences between HC and patients in terms of the magnitude of deviation from normality by performing both a voxel-wise analysis on the thresholded Z-score maps, and on summary measures of deviations. The voxel-wise analysis allows spatial localisation of the significant differences between the two groups, whereas summary measures determine whether it is possible to collapse meaningful information into a few scores per subject which might be useful for clinical applications. The four summary scores analysed were the mean z-score, corresponding to the mean z-score across voxels for each subject, and three measures of extreme deviations from normality, estimated by counting the percentage of voxels with Z-score > 2 (i.e., positive deviations), Z-score < -2 (i.e., negative deviations) or |Z-score| > 2 (i.e., total deviations). Since the clinical datasets included subjects with FEP and chronic psychosis, we ran two-way ANOVAs testing the main effects of cohort (HC, FEP and chronic psychosis) and dataset (FDOPA_01, FDOPA_02, FDOPA_03), and their interaction. The voxel-wise tests were performed using FSL randomise^61^, with 5,000 permutations and considering a cluster significant if p_FWE_ < 0.05^62,63^, corrected for multiple comparisons using the threshold-free cluster enhancement^62^ (TFCE) option. For the significant contrasts, we extracted the mean z-score values from the significant clusters and performed post-hoc tests using the rstatix^58^ library in R 4.2.1. For the tests on the summary metrics, we used the rstatix^58^ library in R 4.2.1.

Lastly, we investigated which biological pathways were driving the spatial pattern of extreme deviations identified in the voxel-wise two-way ANOVA. This was done by running imaging-transcriptomics with the imaging-transcriptomics toolbox^64–66^ on the F-stat map reporting the main effect of the group, and by running GSEA on molecular function, as defined by the GO Molecular Function^67,68^.

### Extending normative model validity to an independent patient cohort

To demonstrate the value of NM analysis when using a clinical cohort without its own group of matched HCs and test the replicability of the results from the previous analyses, we applied the [^18^F]FDOPA PET NM on an independent dataset of patients with psychosis (FDOPA_04). Of note, although this clinical cohort was independent, its scanner effect was modelled in the estimated NM since this dataset was acquired at the same site and using the same scanner as the FDOPA_01 dataset. We estimated the four summary measures (mean Z-score and positive, negative, and total extreme deviations) in this cohort and compared them with the summary measures of the HC of the other datasets (FDOPA 01, 02 and 03). For this analysis, we performed Welch two-sample t-tests using the rstatix^58^ library (R 4.2.1).

### Relationships between deviation scores and clinical symptoms

In datasets where clinical data were available, specifically FDOPA_01 (comprising FEP patients) and FDOPA_04 (comprising a combination of chronic and FEP patients), we tested the hypothesis that NM deviation scores would be associated with symptom severity, as measured using PANSS scores^50^ using voxel-wise comparisons and summary measures. Voxel-wise covariance analysis was conducted utilizing FSL *randomise*^52,53,61^ (independent variables: PANSS scores; dependent variable: Z-score), while Spearman correlations between whole brain summary measures and PANSS scores were computed using rstatix^58^ (R 4.2.1). For the cohort of patients without matched controls (FDOPA_04), we additionally performed a correlation analysis between the individual PANSS scores and the average Z-scores of the statistically significant clusters, identified from the voxel-wise cross-sectional analysis (Spearman correlation, rstatix^58^ library, R 4.2.1).

### Prediction of Treatment Response in Patients

To determine whether the predictive power of FDOPA summary measures outperformed reference analysis (i.e., striatal K_i_), in predicting antipsychotic treatment response for each clinical cohort we constructed ROC curves for all summary measures of the deviation scores (average Z-score, positive, negative, and total extreme deviations) across the whole brain and striatum and ROC curves for the original striatal K_i_ measures. ROC curves were compared using the DeLong test. All analyses were performed using the pROC library^69^ R 4.2.1.

## Results

### Estimation of NM for [^11^C]-(+)-PHNO PET and assessment of the optimal harmonisation strategy

As shown in Figure 1A, the Z-score distributions of the non-harmonised [^11^C]-(+)-PHNO PET data present two distinct scanner-related patterns, which are attenuated after harmonisation with both harmonisation strategies. When comparing the averages of the distributions between scanner types, we found statistically significant differences between the PET/CT and PET/MR scanners both in the unharmonized data (t=2.46, p<0.01) and in the data harmonised with smoothing (t=2.85, p<0.01), while there were no scanner-related differences in the data harmonised with Combat (t=0.71, p=n.s.). In terms of standard deviation, we found significant differences between scanner types in the non-harmonised data (t=-14.59, p<0.001) and residual differences in the data harmonised with Combat (t=-2.47, p<0.01), while no differences between scanner types were found in the data harmonised with smoothing (t=-0.40, p=n.s.).

**Figure 1.**
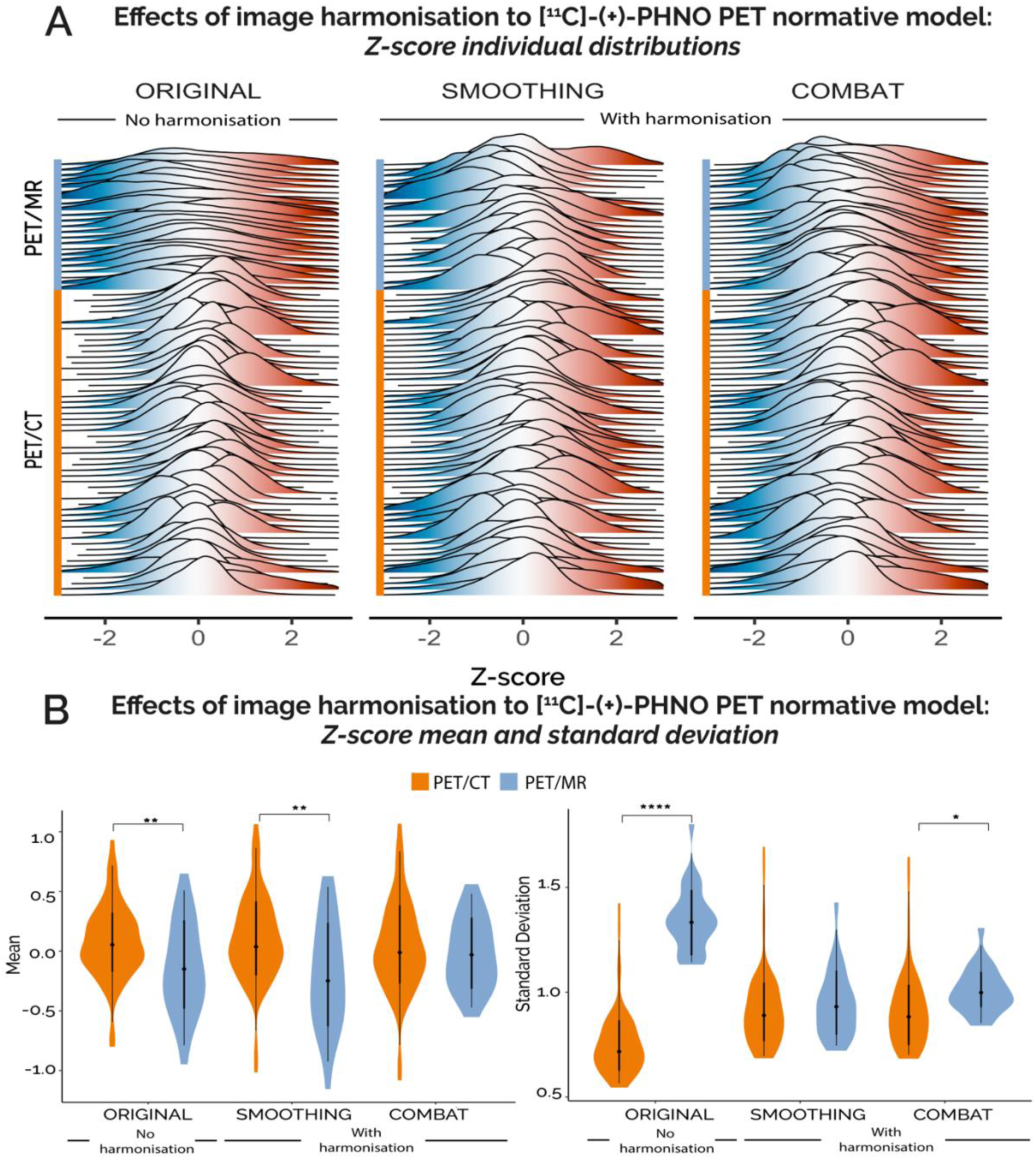
Harmonisation results of [^11^C]-(+)-PHNO PET normative modelling. (A) Single-subject, whole-brain distributions of Z-scores estimated in the data before harmonisation (original data, on the left) and after spatial smoothing (in the centre) and Combat (on the right) harmonisation methods. This dataset combines scans of healthy controls acquired with either PET/CT (N=54) or PET/MR (N=23) scanners. (B) Welch two-sample t-tests comparing the first statistical moments (mean and standard deviation of the single-subject Z-score distributions between scanner types for each harmonisation modality (no harmonisation, harmonisation with spatial smoothing and harmonisation with Combat). Orange violin plots refer to PET/CT scans while blue violin plots refer to PET/MR scans. Asterisks indicates significance. * indicates p<0.05, ** indicates p<0.01 and **** indicates p<<0.001.

When analysing between-scanner differences at the regional level (Supplementary figure 1), all harmonisation methods showed significant differences in the number of extreme deviations between scanner types (p<0.01), except Combat for the negative extreme deviations. Overall, Combat returned the smallest deltas, hence better harmonisation performances, for both positive extreme deviations (No harmonisation: Δ_MR-CT_ = 4.30±2.21%; Smoothing: Δ_MR-CT_ = -1.03±1.39%; Combat: Δ_MR-CT_ = 0.44±1.04) and negative extreme deviations (Original: Δ_MR-CT_ = 6.02±2.30%; Smoothing: Δ_MR-CT_ = 2.06±2.10%; Combat: Δ_MR-CT_=0.06±0.97%) (Supplementary Figure 1).

### Validation of the optimal harmonisation strategy on [^18^F]FDOPA data

After harmonising [^18^F]FDOPA data with Combat, we estimated the voxel specific NMs, which explained up to 15% of the data variability although they do not converge to a solution for approximately 56% of the grey matter voxels (i.e., EXPV < 0; Supplementary Figure 2). All subsequent results will refer to NM data thresholded with the 3% variance mask, although the same analyses were repeated for the other masks (Supplementary Tables 1-12).

No clear scanner effects were apparent from the individual distributions of Z-scores in HC (Supplementary Figure 3). Similar to [^11^C]-(+)-PHNO PET NM, for FDOPA no significant scanner-related effect was found in terms of mean z-score distribution (F=0.12, p=n.s.), while a significant residual effect was found for the standard deviation (F=3.38, p<0.05). Indicating that the harmonisation strategy was effectively applied to FDOPA.

### NM application to [^18^F]FDOPA PET in psychosis

Grouped distributions of HC and patients showed small differences in the mean (|Δ_μ_^HC-PAT^| = 0.14), and a greater difference in terms of the standard deviation (|Δ_σ_^HC-PAT^| = 0.28), consistently higher for patients in both measures. The number of extremely deviating voxels (in percentage to the total voxels in the analysis mask), above the level of chance, shows increased RR for patients with all extreme deviation scores (RR_pos_=2.19, RR_neg_=1.92 and RR_tot_=2.60 for positive-, negative-, and total-extreme deviations respectively) (Supplementary Figure 2).

We hypothesised that the HC group would exhibit a relatively low percentage of individuals with extreme deviations, while conversely, patients would demonstrate a higher overlap in the percentage of individuals with extreme deviations from normality in specific brain regions. While HC reported very low percentages of individuals with overlapping extreme deviations (Figure 2), which aligns with the NM hypothesis for the reference cohort, patients exhibited distinct spatial patterns of extreme deviations, primarily in the cortex. These deviations were prominently present in the precentral and frontal gyri. However, it is important to emphasise that this pattern of extreme deviations also showed substantial sample heterogeneity, as the highest percentage of patients with comparable extreme deviations did not exceed 20%.

**Figure 2.**
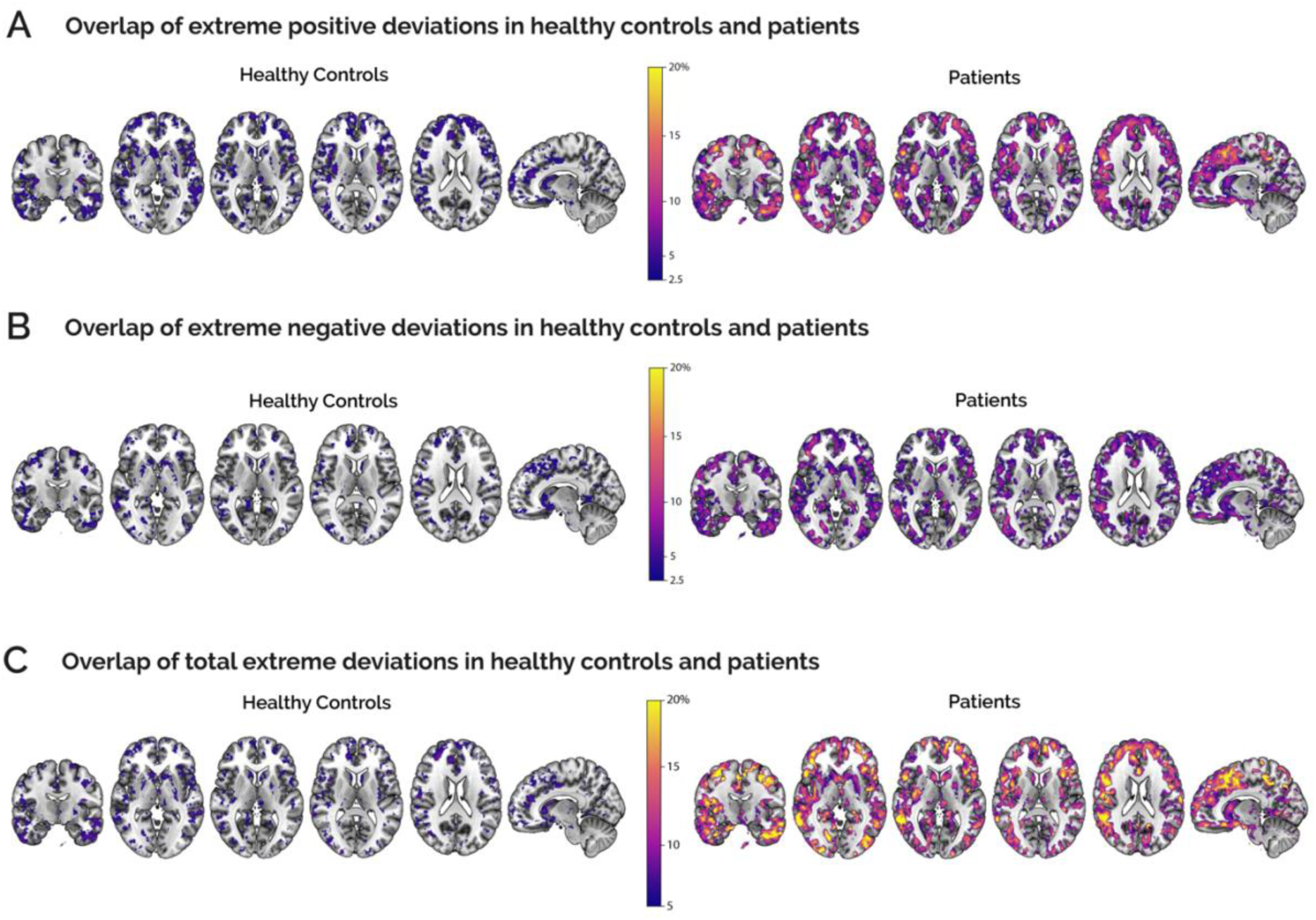
Overlap maps of extreme deviations in HC and patients. Each map reports for each voxel the percentage of subjects with co-localised positive (top row), negative (middle row) and total (bottom row) extreme deviations. Controls (left) and psychosis patients (right) are reported separately. The figure is set at a threshold above the level of chance (2.5% for positive and negative deviations and 5% for total deviations). All overlap maps are superimposed on a standard MNI152 structural template.

The voxel-wise two-way ANOVA performed on the non-thresholded Z-score maps revealed significant interactions between group and dataset (F_peak_=21.71, p_peak_<0.001 FWE corrected). This interaction spanned the temporal lobe, the medial- and superior-orbitofrontal, and the frontal lobe. The post hoc tests to evaluate between-group differences within each dataset highlighted greater positive deviations in the FEP as compared to the HC (FDOPA_01, t =4.92, p<<0.001) and lower negative deviations in chronic patients as compared to HC in both FDOPA_02 (t =2.22, p <0.05) and FDOPA_03 (t =4.62, p <0.001).

We also found a group effect in the two-way ANOVA performed on the total extreme deviation maps (F_peak_=34.36, p<0.001 FWE corrected) (Figure 3), exhibiting an increase in patients. This effect was localised in the left frontal orbital cortex, frontal pole, and inferior temporal cortex. Of note, only clusters including more than 50 voxels are reported.

**Figure 3.**
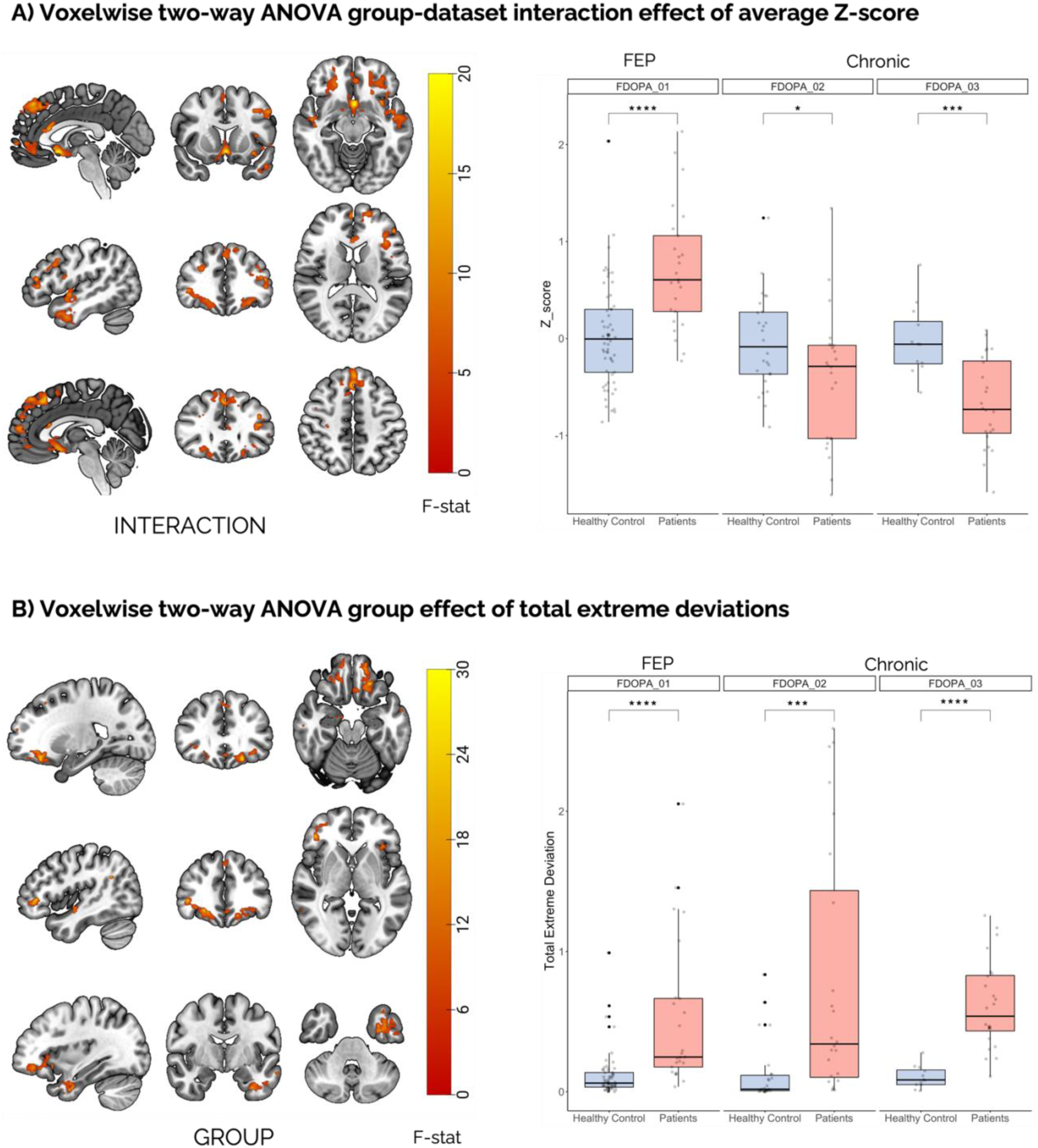
Voxel-wise ANOVA analysis of deviation scores, expressed as mean Z-score and total extreme deviations, for the [^18^F]FDOPA Normative Model. (A) Mean Z-score significant clusters (left) and post-hoc analyses, grouped by dataset, (right) between healthy controls and patients. (B) Total extreme deviations significant clusters (left) and post-hoc analyses, grouped by dataset, (right) between healthy controls and patients. * indicates p<0.05, *** indicates p<0.001 and **** indicates p<<0.001.

The two-way ANOVAs conducted on the summary measures of the mean z-score and extreme positive, negative, and total deviations revealed a significant interaction between group and dataset for the mean Z-score (F = 8.43, p<0.001; Figure 4A), extreme positive deviations (F=5.90, p<0.01; Figure 4B) and extreme negative deviations (F=5.89, p<0.01, Figure 4C). Post hoc analyses showed an increase, in average Z-score in FEP (t =3.08, p<0.01), and a decrease in chronic patients (t =-3.05, p<0.01) compared to HCs (Supplementary Table 5). Similarly, FEP was associated with an increase in extreme positive deviations compared to HCs (t =3.23, p<0.01, Supplementary Table 6), whereas there was an increase of extreme negative deviations in chronic patients compared to HCs (FDOPA_02, t= 2.67, p<0.05; FDOPA_03, t =3.36, p<0.01; Supplementary Table 7). The total extreme deviations, which combine positive and negative extreme values, did not exhibit a significant interaction between the group and dataset, although it did indicate a significant effect of the group (F=37.52, p<<0.001, Figure 4D, Supplementary Table 8), showing an increase in patients compared to HC.

**Figure 4.**
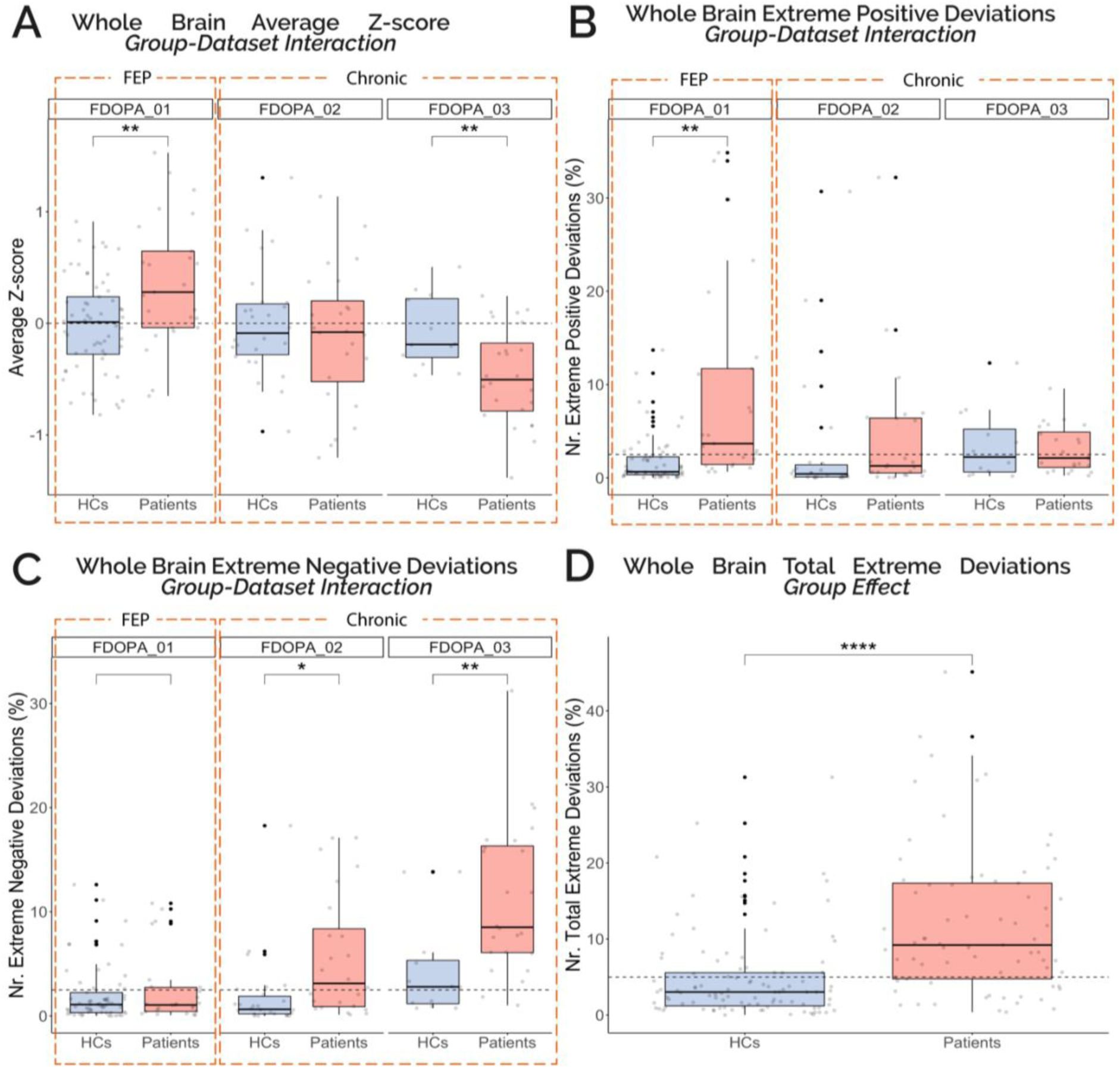
ANOVA analysis of individual deviation scores, expressed as mean Z-score and positive, negative, and total extreme deviations, for the [^18^F]FDOPA Normative Model. (A) Mean Z-score differences between healthy controls and patients grouped by dataset. (B) Difference of extreme positive deviations between healthy controls and patients grouped by dataset (C) Difference of extreme negative deviations between healthy controls and patients grouped by dataset. (D) Difference of total extreme deviations between group. Patients are composed of a mixture of first episode (FDOPA_01) and chronic psychosis (FDOPA_02, FDOPA_03), depending on the dataset. Asterisks indicates significance, * indicates p<0.05, ** indicates p<0.01 and **** indicates p<<0.001.

The imaging transcriptomic analysis conducted on the F-stat maps resulting from the significant group effect on the total extreme deviations, tested with two-way ANOVA testing and representing abnormalities in patients regardless of the duration of psychosis, revealed several notable enriched gene sets. These included “structural constituent of ribosome” (NES=2.49, p_FDR_<0.05), “neurotransmitter receptor activity” (NES=2.18, p_FDR_<0.05) and “glutamate receptor activity” (NES=2.04, p_FDR_<0.05), as determined by the GO molecular function^67,68^.

### Extending normative model validity to an independent patient cohort

When visually compared with the HC of the independent datasets in terms of the percentage of individuals with extreme deviations (positive, negative, and total) within the group, the pattern of extreme deviations of the FDOPA_04 dataset is consistent with that of the datasets of patients in FDOPA_01, FDOPA_02 and FDOPA_03 (Supplementary Figures 5-7).

In terms of differences in the summary measures (mean z-score, and positive, negative, and total extreme deviations) between FDOPA_04 patients and HC, we found significant increases in patients in terms of mean Z-score (t=-3.65, p<0.001), extreme positive deviation (t=-5.52, p<0.001) and total extreme deviations (t=-5.93, p<0.001). No significant differences were found in terms of negative extreme deviations (t=-1.14, p=n.s.).

### Correlation between imaging-based deviations from normality and clinical symptoms

We tested the hypothesis that there would be an association between the significant clusters from the voxel-wise two-way ANOVA performed on the extreme deviation maps and PANSS scores in both the FDOPA_01 (FEP) and FDOPA_04 (chronic schizophrenia) datasets. There were no significant correlations in the FDOPA_01 dataset. In the FDOPA_04 dataset, the mean Z-scores and total extreme deviations were significantly correlated with clinical scores (Table 2). Three individual clusters in FDOPA_04 (with n_voxels_≥50) survived correction for multiple comparisons, showing significant correlations with total PANSS scores (Table 3).

**Table 2.** Correlations between significant voxel-wise clusters and PANSS scores. The table shows Spearman correlation’s rho and relative p-value for significant clusters resulting from voxel-wise two-way ANOVA (average Z-score, and |Z|>2). Correlations are separated for FDOPA_01 (FEP patients, N=25) and FDOPA_04 (chronic and FEP patients, N=36). * indicates p<0.05, ** indicates p<0.01

|  | PANSS Positive |  | PANSS Negative |  | PANSS General |  | PANSS Total |  |
| --- | --- | --- | --- | --- | --- | --- | --- | --- |
|  | FDOPA_01 | FDOPA_04 | FDOPA_01 | FDOPA_04 | FDOPA_01 | FDOPA_04 | FDOPA_01 | FDOPA_04 |
| Average Z-score | 0.38 | 0.36* | 0.38 | 0.38* | -0.06 | 0.41* | 0.36 | 0.43** |
| Extreme total deviations | 0.32 | 0.36* | 0.31 | 0.34* | -0.09 | 0.42* | 0.25 | 0.44** |

**Table 3.**
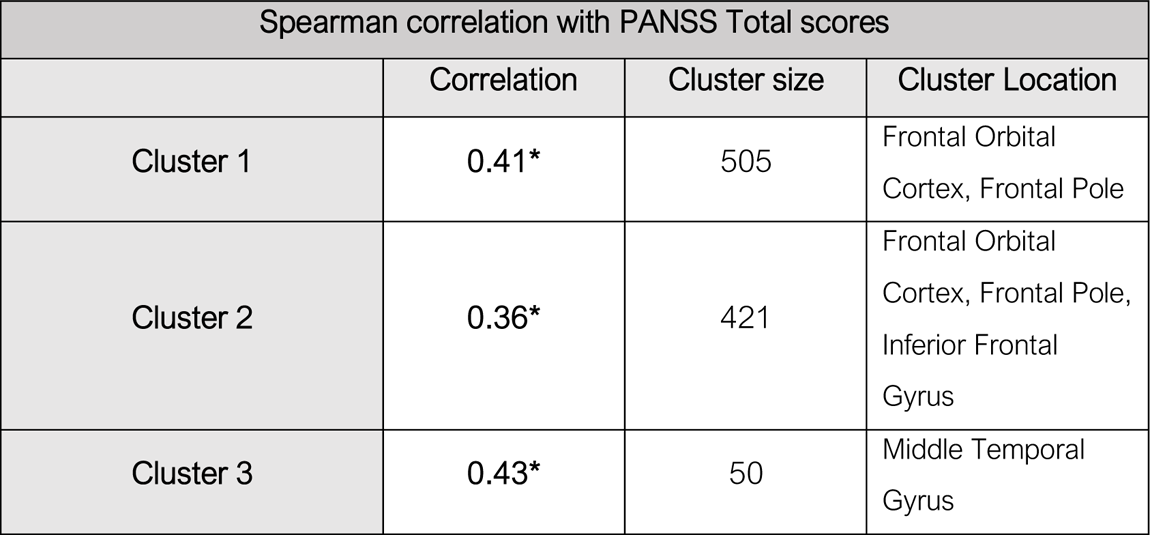
Correlation analysis of PANSS total scores and individual voxel-wise clusters. Table shows Spearman correlation’s rho and relative p-value for significant clusters between total extreme deviations and PANSS Total Scores in FDOPA_04 (chronic patients), resulting from voxel-wise two-way ANOVA (|Z|>2). Cluster locations were obtained using FSL atlasquery from the Harvard-Oxford Cortical Atlas. * indicates p<0.05.

Correlation of the summary deviations and PANSS scores revealed a significant correlation between total extreme deviations and PANSS negative in FEP patients (i.e., FDOPA_01). The analysis of FDOPA_04 reveals significant correlations between extreme positive deviations and PANSS general scores and between total extreme deviations and all PANSS scores (Table 3).

**Table 4.**
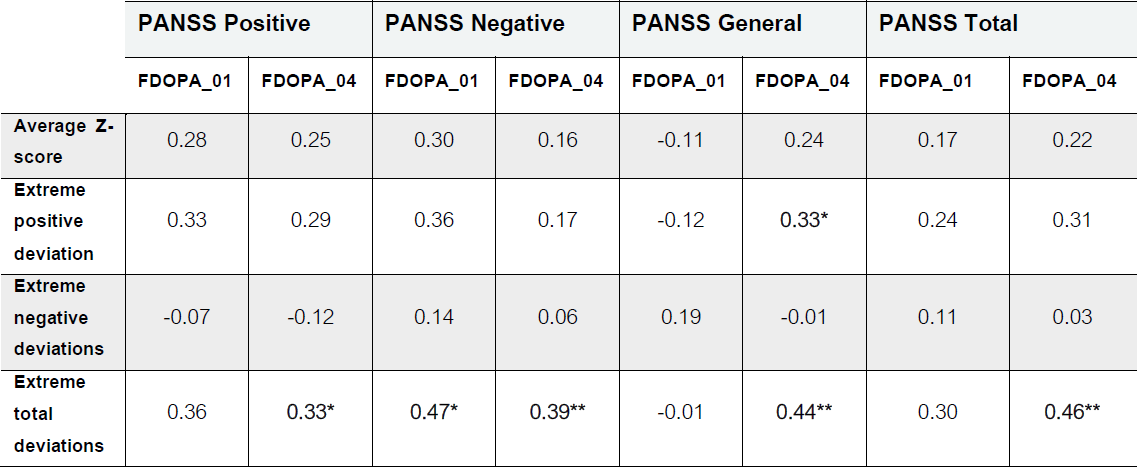
Correlation analysis of PANSS scores and summary measures. Table shows Spearman correlation’s rho and relative p-value for whole brain summary measures (average Z-score, Z>2, Z<-2, |Z|>2). Correlations are separated for FDOPA_01 (FEP patients, N=25) and FDOPA_04 (chronic and FEP patients, N=36). * indicates p<0.05, ** indicates p<0.01

### Treatment Response in Patients

The analysis on the summary mean Z-score to evaluate patients’ response to standard neuroleptics replicated the performance of the reference analysis (i.e., K_i_^cer^) and showed no significant difference between the two methods (as measured by the DeLong test) when focusing on the striatum only (Figure 5A). Here we obtained acceptable performances for FDOPA_02 (AUC_str_=0.77, AUC_Ki_=0.79)^70^, and excellent performances for FDOPA_01 (AUC_str_=0.83, AUC_Ki_=0.83) and FDOPA_03 (AUC_str_=0.83, AUC_Ki_=0.74)^70^. However, when extending the analysis to the summary measures estimated at the whole brain, we did not reach acceptable classifications except for FDOPA_02 (AUC=0.70, Figure 5A). Classification of treatment response measured with whole-brain deviation scores pointed at acceptable^70^ performances only for FDOPA_02 in terms of negative extreme deviations (AUC=0.72, Figure 5C).

**Figure 5.**
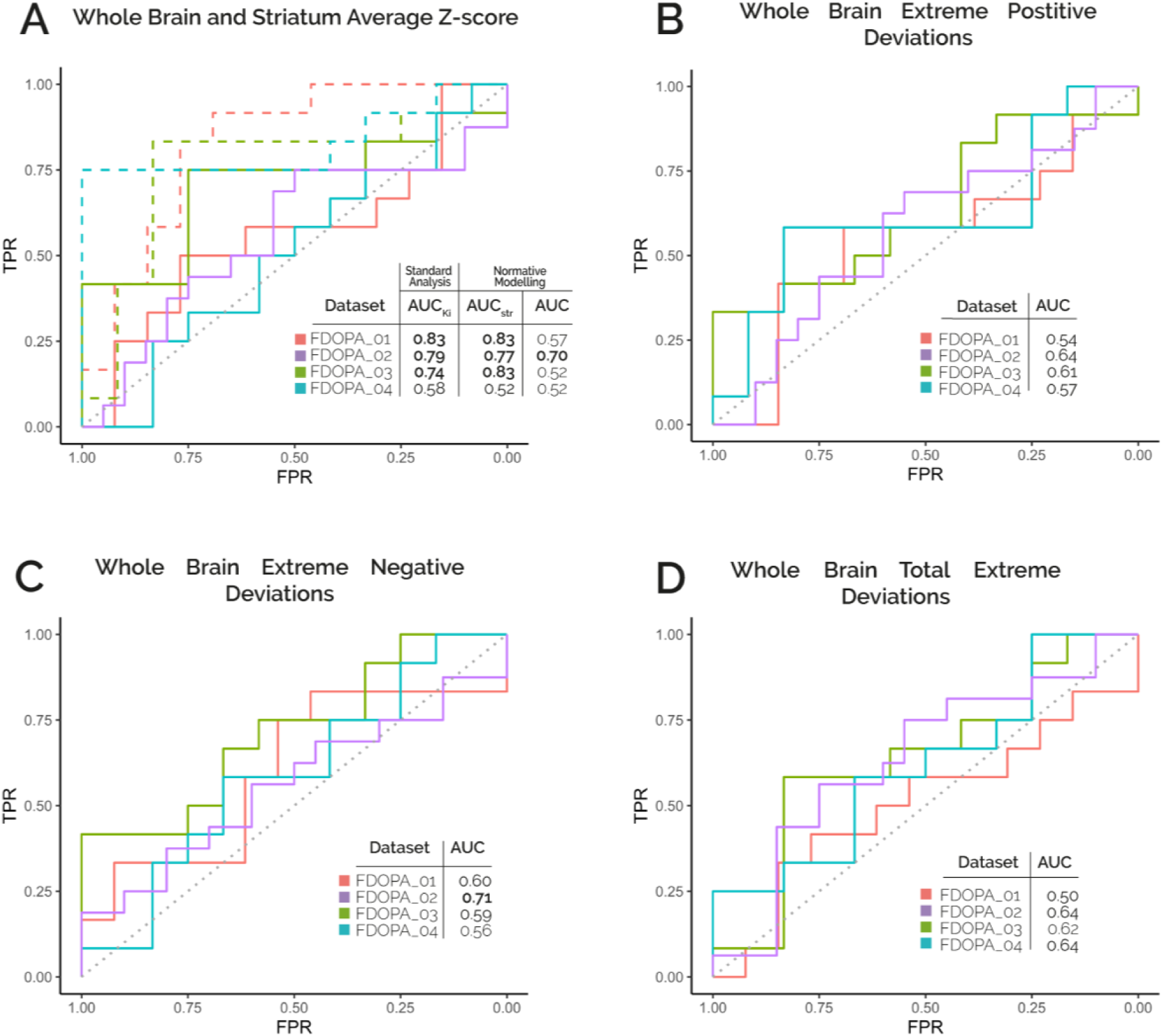
Clinical [^18^F]FDOPA Normative Model Treatment Response. ROC curves for the classification of treatment response (Responders vs Non-Responders/Resistant) using whole brain (solid lines) or striatal (dashed lines) individual average Z-score (A), whole brain positive extreme deviations (B), negative extreme deviations (C) and total extreme deviations (D) for the different clinical datasets. Dashed diagonal indicates the level of chance in all plots. All scores refer to [^18^F]FDOPA patient data after Combat harmonization.

## Discussion

This study applied normative modelling (NM) in PET imaging of the dopamine system in psychosis using several existing datasets. Two dopaminergic radiotracers ([^11^C]-(+)- PHNO for D_2/3_ receptor and [^18^F]FDOPA for dopamine synthesis) were used to construct normative models from reference healthy controls and applied to patients with psychosis. We demonstrate that NM is capable of explaining dopamine function variability arising from demographic factors and show that psychosis is associated with a differential pattern of extreme deviations. [^18^F]FDOPA PET showed a high degree of patient heterogeneity, particularly in extra-striatal brain areas. In the striatum, deviation scores were able to classify treatment response, although with similar performance to standard kinetic modelling analysis. Overall, NM provides novel quantitative insights into the brain dopamine system in healthy individuals and people with psychosis, taking advantage of existing normative data built from multiple neuroimaging studies.

### Considerations on NM applied to molecular neuroimaging

In our study, we first confirmed the necessity of data harmonisation to compile multi-centre, multi-scanner PET datasets with the required sample size for constructing a normative model of healthy cohorts. Our results indicated that Combat was the best harmonisation method to remove scanner and site effects. This finding aligns with the neuroimaging literature, where model-based harmonization methods are the preferred choices in PET^71^ and other modalities^32,33^. It’s worth noting that Combat assumes linearity in the scanner’s effect, which may not hold true for tomographs with different geometries, axial fields of view, and gamma detectors. Recently, non-linear versions of Combat methods (e.g., Combat-GAM^72^) have been introduced, but they would require further testing with PET tracers exhibiting different kinetic properties. Regardless of the type of harmonization statistics used, our proposed approach relies on the existence of a group of healthy controls (HC) for scanner calibration. This results in a non-generalizable model without HC data. One solution, proposed in other neuroimaging modalities^27,28,28,73^, could be to incorporate the scanner directly into the normative model as a random effect using hierarchical modelling. While appealing for modelling flexibility, this methodology is burdened by extreme computational times (reported to be up to ∼24 hours per model^27^), making it unfeasible for our application (i.e., ∼130K voxel-wise models).

Our normative model was able to explain a substantial amount of dopamine function variability (∼30% in D_2/3_ receptor density, as measured by PHNO, and ∼15% in dopamine synthesis capacity, as measured by FDOPA) using simple demographic factors from the population. This level of variance is lower than that reported in other modalities (40% for sMRI^35^, 29% for diffusion MRI^74^), which may relate to measurement of brain functional processes rather than structure. To allow generalizability, the covariates we selected were limited to general demographic factors (age and sex for [^18^F]FDOPA; age, sex, and BMI for [^11^C]-(+)-PHNO) were not specific to psychosis. Although the addition of more diagnosis-specific covariates and/or additional experimental factors (e.g., genetic traits), may have improved the normative model’s performance, the use of ancillary measures (e.g., genetic tests or clinical assessments) might limit its potential clinical utility due to the necessity of having a PET scan and additional testing. Our model is therefore disease-agnostic.

### Considerations on NM applied to FDOPA PET imaging and patients with psychosis

The application of an FDOPA NM to data acquired patients with psychosis provided several novel pathophysiological insights. Firstly, we observed that patients exhibit a higher percentage of extreme deviations than HCs. This observation is further supported by the elevated risk ratio (RR) for both positive and negative extreme deviations in patients compared to controls. Surprisingly, summary Z-score measures (calculated as individual whole-brain summary scores) exhibit a differential pattern consistent with the chronicity of the disease (i.e., first-episode psychosis – FEP, or chronic patients).

We observed that while there is a substantial overall difference due to the disease (as indicated by the ANOVA analysis of total extreme deviations), the pattern of positive and negative extreme deviations varies in different stages of the disease. For example, FEP patients show a higher number of positive extreme deviations than healthy controls and chronic patients, while the latter show a higher number of negative extreme deviations than controls and FEP patients. This pattern is also evident in the average distribution scores and is confirmed by significant voxel-wise analyses (average Z-score and total extreme deviations).

Plausible explanations for this differential pattern are the effect of medications acting on the dopaminergic system, particularly D_2/3_ receptors, or grey matter reductions in chronic patients when compared to FEP^75^. Long-term antipsychotic treatment may up-regulate D_2/3_ receptors, which could potentially impact presynaptic dopamine synthesis capacity also through pre-synaptic autoregulatory mechanisms.

With the data available to our study, we cannot establish whether this differential pattern between FEP and chronic patients is attributed to variances in disease duration or is linked to medication effects. It is worth noting that each patient’s experience with the disease is unique, and ideally, they should not be grouped with other patients for analytical purposes but rather considered as individual "clusters"^76^. Nevertheless, by employing NM, it would be feasible (provided the requisite data is available) to compare individual patients with consideration to their clinical and disease history.

Another interesting finding from our analyses is the substantial heterogeneity among patients, even though there is a consistent effect in all patients. This heterogeneity is particularly noticeable in analysis of the overlap of extreme deviations, where at most, 20% of patients co-localize in the same voxels. The co-localization of extreme deviations is more pronounced in patients compared to healthy controls (with a maximum expected overlap of 5%), indicating a common alteration of [^18^F]FDOPA PET signal in psychosis. These areas of extreme deviations are consistent with previous findings on NM applied to structural MRI, showing a maximum degree of overlap of 5-10%^19,20^. Future studies should explore of interlinks between structural and molecular changes at the individual level. Interestingly, [^18^F]FDOPA PET co-localizations of extreme deviations in psychosis occur in extra-striatal brain areas, contrary to the focus of most [^18^F]FDOPA studies on striatal areas^38–47,49^. Studies which have examined the cortical signal of [^18^F]FDOPA suggest elevations in dopamine synthesis capacity in cortical areas (e.g., posterior cingulate^77^, medial prefrontal cortex^78^) or correlations between dopamine synthesis capacity in cortical areas and clinical symptom severity (e.g., a positive correlation between PANSS positive scores and K_i_ in the right temporal cortex^79^). These findings raise questions about alterations in aromatic L-amino acid decarboxylase (AADC) and whether patients have an unresponsive feedback mechanism of tyrosine hydroxylase, which may not down-regulate as in normal populations^77^. Early animal studies suggest that the [^18^F]FDOPA signal combines metabolic activity related to dopamine synthesis, storage, and metabolism^80^. Our imaging-transcriptomics analysis partially confirms these findings pointing towards an association between genetic pathways related to metabolism and [^18^F]FDOPA deviations measured in psychosis. All these findings suggest that cortical signal of [^18^F]FDOPA might be associated with increased cellular transport and/or protein metabolism, supporting the application of this biomarker in neuro-oncology^81^. Further studies are required to fully characterise the nature of the [^18^F]FDOPA signal in the human cortex, establishing the contributions of dopamine function (e.g., neuron density and changes in dopaminergic-mediated neuronal firing), metabolism and cellular transport to the overall signal. Lastly, there are genetic variants that are well-known to influence the dopamine system (e.g. 22q11.2 deletion/duplication^40^ and COMT Val^158^Met polymorphism^82^) or that have been associated with the [^18^F]FDOPA PET signal (e.g. AS3MT/BORCS7 genetic variant^83^). These factors could drive some of the functional differences observed between patients and HCs and increase the inter-individual explained variance if included in the NM. Further analyses in a larger sample with full genetic information are warranted.

In our study, we explored the feasibility of investigating dopaminergic alterations of [^18^F]FDOPA without acquiring a matched set of healthy controls. With the analysis of the patient-only cohort (i.e., FDOPA_04) and its comparison to the whole normative reference (i.e., all HCs) we obtained similar results as those in the other clinical datasets which had matched HCs in the normative reference, including the spatial pattern of the extreme deviations. This finding is particularly relevant, as routine clinical applications typically do not acquire matched controls for patients. The NM approach circumvents this issue by using a normative reference, accounting for covariates for comparison. Furthermore, the deviation scores of FDOPA_04, especially total deviations, showed significant correlations with PANSS symptoms, both as whole-brain summary scores and significant cluster measures.

Interestingly, by analysing individual clusters within the cluster signal, we found that the clusters correlating with total PANSS symptoms were in the frontal and temporal gyri. However, no significant correlations were observed in the smaller FDOPA_01 dataset, which may relate to lower statistical power. This framework, along with automatic standardised frameworks for kinetic modelling of [^18^F]FDOPA^49^, represents a step forward in making PET parametric imaging a quantitative companion tool for clinical management of psychosis patients.

Finally, we analysed the potential of using NM-derived scores for classifying treatment response in patients (i.e., those who will respond to treatment at follow-up based on baseline scores) and compared these findings to previously published results^38^. Interestingly, when focusing on striatum NM, we replicated the prediction performances of the reference standard^38^, while poor classification performances were obtained when extending the method to the whole brain. This outcome aligns with expectations, as the molecular target of action of both antipsychotic medications is striatal dopamine D_2/3_ receptors^84,85^. In fact, the striatal average Z-scores (as defined by the Hammersmith atlas and where the model converges) were highly correlated with K_i_ estimates in all patient datasets (0.93<R<0.97, p<0.001, Supplementary Figure 8), providing thus an alternative view of the same information. In line with this, the distributions of extreme positive and total extreme deviations in the striatal region revealed group differences, indicating an elevation in the dopamine signal (Supplementary Figure 9).

### Limitations

This work is subject to several limitations. Firstly, the modelling process is influenced by the harmonization method and may not account for non-linearities within the system, either in harmonization or modelling. Despite our efforts to harmonize data from different clinical sites, residual differences still exist in the datasets when assessed with statistics of the second order or above (e.g., skewness, kurtosis). Secondly, applying NM at the voxel level can result in underperformance of the model due to high noise and poor signal. Consequently, there are a considerable number of voxels where the model fails to converge. This issue could potentially be addressed by adjusting the MCMC settings or by incorporating a more extensive set of covariates to describe the neuroimaging target variable. While NM mapping provides a topological description of patient deviation, region-based modelling remains a valid compromise between spatial resolution and method performance. Additionally, the deviation scores might be sensitive to motion^86^, however when assessing for our data there were no significant correlations in HCs nor in patients (Supplementary Figure 10). Moreover, the heterogeneity of the clinical sample used in the analysis and the presence of missing clinical information for some individuals in the cohort are likely to have affected statistical power. Lastly, our result could be influenced by the choice of EXPV threshold used for masking, although similar results were obtained for the other threshold tested (Supplementary Tables 1-12).

### Conclusions

In conclusion, the NM framework can be successfully applied to molecular neuroimaging (i.e., PET and SPECT) after proper harmonisation of scanner effects. Moreover, with the NM model, we can assess a differential pattern of deviations likely attributable to the chronicity of the disease and compare a patient-only cohort to the normative reference to gain mechanistic insights and advance toward a quantitative and biological understanding of psychosis. Additionally, we are able to replicate the findings of traditional cross-sectional studies and performances with standard analytical approaches. While the focus of this work was on the presynaptic dopaminergic system (as measured by [^18^F]FDOPA) and on the post-synaptic dopaminergic system (as measured by [^11^C]-(+)- PHNO) in the context of psychosis, we believe that after selecting appropriate covariates, this methodology can be applied to any molecular target measured by PET or SPECT neuroimaging.

## Data Availability

The data that support the findings of this study are available from The NeurOimaging DatabasE (NODE) repository (https://maudsleybrc.nihr.ac.uk/research/precision-psychiatry/neuroimaging/neuroimaging-database-node/) but restrictions apply to the availability of these data, which were used under license for the current study, and so are not publicly available. Data are however available from the authors upon reasonable request and with permission by the data controller institutions, by contacting the support team or the author Dr. Giovanna Nordio.

## Acknowledgements

This study represents independent research funded by the NIHR Maudsley Biomedical Research Centre at South London and Maudsley NHS Foundation Trust and King’s College London. The views expressed are those of the author(s) and not necessarily those of the NIHR or the Department of Health and Social Care. A.G. is supported by the KCL-funded CDT in Data-Driven Health; this represents independent research partly funded by the NIHR Maudsley’s Biomedical Research Centre (BRC) at the South London and Maudsley NHS Foundation Trust and partly funded by GSK. D.M., G.N., R.E., O.D., F.T., and S.C.R.W. are supported by the NIHR Maudsley’s Biomedical Research Centre at the South London and Maudsley NHS Trust. MV is supported by the Italian National Centre for HPC, BIG DATA AND QUANTUM COMPUTING (Project no. CN00000013 CN1), the PNR Italian National Grant DIGITAL LIFELONG PREVENTION (Project no PNC0000002_DARE), and by Wellcome Trust Digital Award (no. 215747/Z/19/Z). M.d.G. is an employee of GSK, GSK had no role in the design of this study.

## Conflict of Interests

M.d.G. is an employee of GSK, GSK had no role in the design of this study. R.A.M. has received speaker/consultancy fees from Karuna, Janssen, Boehringer Ingelheim, and Otsuka, and co-directs a company that designs digital resources to support treatment of mental illness. F.B. has received consulting fees from Petalouda Therapeutics and has been an employee at Compass Pathways. AE has received consulting fees from Leal Therapeutics.

## Supplementary Information

Supplementary information is available at MP’s website.

## Notes

### Author Declarations

All the research protocols for data acquisitions were approved by local ethics committees and institutional revision boards including the Institute of Psychiatry, King's College, London, England, research ethics committee; the South London and Maudsley/Institute of Psychiatry NHS Trust, London-West London & GTAC Research Ethics Committee; the Administration of Radioactive Substances Advisory Committee (ARSAC); the Hammersmith Research Ethics Committee; the East of England-Cambridge East NHS Research Ethics Committee; Seoul National University Hospital, Seoul, Korea.

