## Supplementary Materials for "Investigating Dopaminergic Abnormalities in Psychosis with Normative Modelling and Multisite Molecular Neuroimaging"

FDOPA PET imaging working group (in alphabetical order):

Ilinca Angelescu[1,2], Michael Bloomfield[3], Ilaria Bonoldi[1], Faith Borgan[1], Enrico D'Ambrosio[1,4], Tarik Dahoun[5,6,7], Arsime Demjaha[1], Alice Egerton[1], Sameer Jauhar[1,5,6], Stephen Kaar[1,5,8,9], Euitae Kim[10,11,12], Seoyoung Kim[10], James MacCabe[1], Julian Matthews[13], Robert McCutcheon[1,14,15], Philip McGuire[14], Chiara Nosarti[16,17], Maria Rogdaki[1], Grazia Rutigliano[1,5], Peter S Talbot[18], Luke Vano[1,5]

[1] Department of Psychosis Studies, Institute of Psychiatry, Psychology & Neuroscience, King's College London, London, United Kingdom

[2] Max Planck UCL Centre for Computational Psychiatry and Ageing Research, Institute of Neurology, London, UK

[3] Division of Psychiatry, Faculty of Brain Sciences, University College of London, London, UK
[4] Group of Psychiatric Neuroscience, Department of Translational Biomedicine and Neuroscience, University of Bari "Aldo Moro", Bari, Italy
[5] Psychiatric Imaging Group, MRC London Institute of Medical Sciences, Hammersmith Hospital, Imperial College London, London, UK
[6] Institute of Clinical Sciences, Faculty of Medicine, Imperial College, Imperial College London, London, UK
[7] Department of Psychiatry, Warneford Hospital, University of Oxford, Oxford, UK
[8] Institute of Clinical Sciences (ICS), Faculty of Medicine, Imperial College London, UK
[9] South London and Maudsley NHS Foundation Trust, London, UK
[10] Department of Psychiatry, Seoul National University Bundang Hospital, Gyeonggi-do, Republic of Korea
[11] Department of Psychiatry, College of Medicine, Seoul National University, Seoul, Republic of Korea [12] Department of Brain & Cognitive Sciences, College of Natural Sciences, Seoul National University, Seoul, Republic of Korea
[13] Division of Neuroscience and Experimental Psychology, School of Biological Sciences, Faculty of Biology, Medicine and Health, University of Manchester, Manchester, UK
[14] Department of Psychiatry, Warneford Hospital, University of Oxford, Oxford, OX3 7JX, UK
[15] Oxford Health NHS Foundation Trust, Oxford, OX3 7JX, UK
[16] Department of Child and Adolescent Psychiatry, Institute of Psychiatry, Psychology & Neurosciences, King's College London, London, UK
[17] Centre for the Developing Brain, Division of Imaging Sciences & Biomedical Engineering, King's College London, London, UK
[18] Division of Neuroscience and Experimental Psychology, School of Biological Sciences, Faculty of Biology, Medicine and Health, University of Manchester, Manchester, UK

### SUPPLEMENTARY

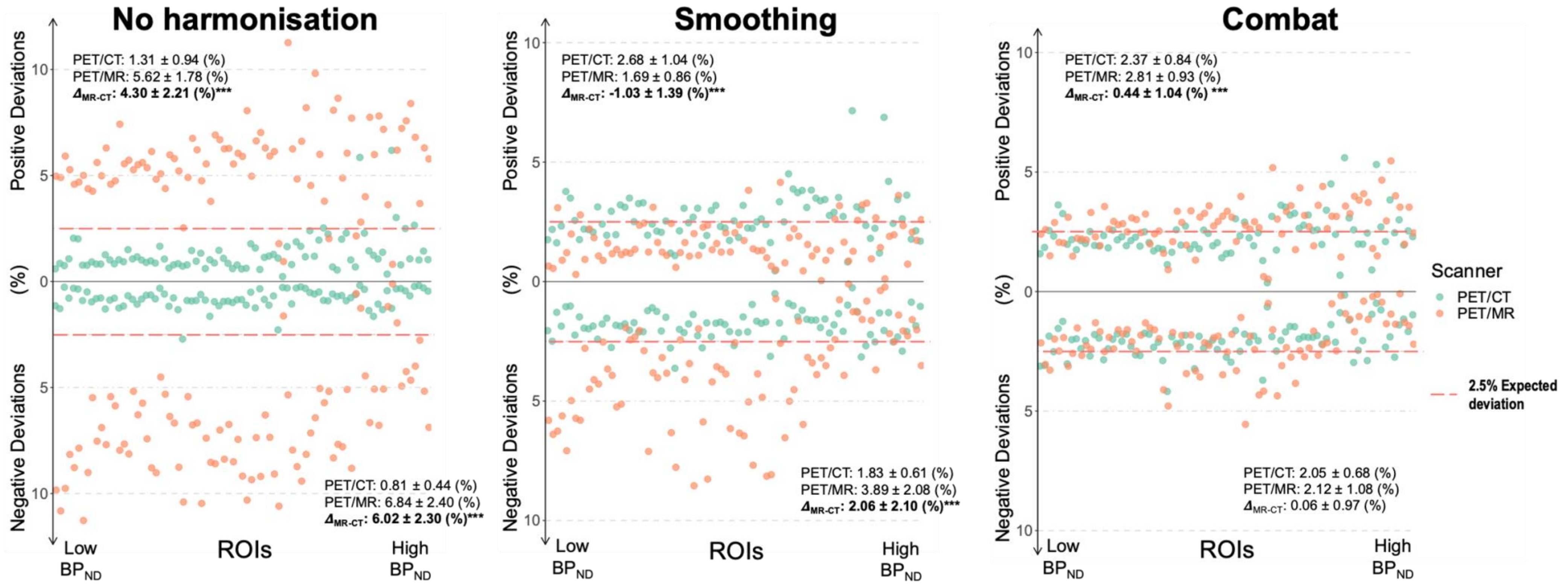


**Supplementary figure 1: [11C]-(+)-PHNO scanner difference of regional extreme deviations between harmonisation methods**. Figure shows extreme positive (Z>2) and negative (Z<-2) deviations of [11C]-(+)-PHNO normative model in regions defined by the Hammersmith atlas. X axis represents ROIs ordered from the lowest to highest binding (i.e., $BP_{ND}$) of the tracer. Green dots represent scans acquired with the PET/CT scanner while orange represent scans acquired with the PET/MR. Dashed line represent the expected fraction of voxels exceeding the selected threshold for extreme deviation (|Z|>2).

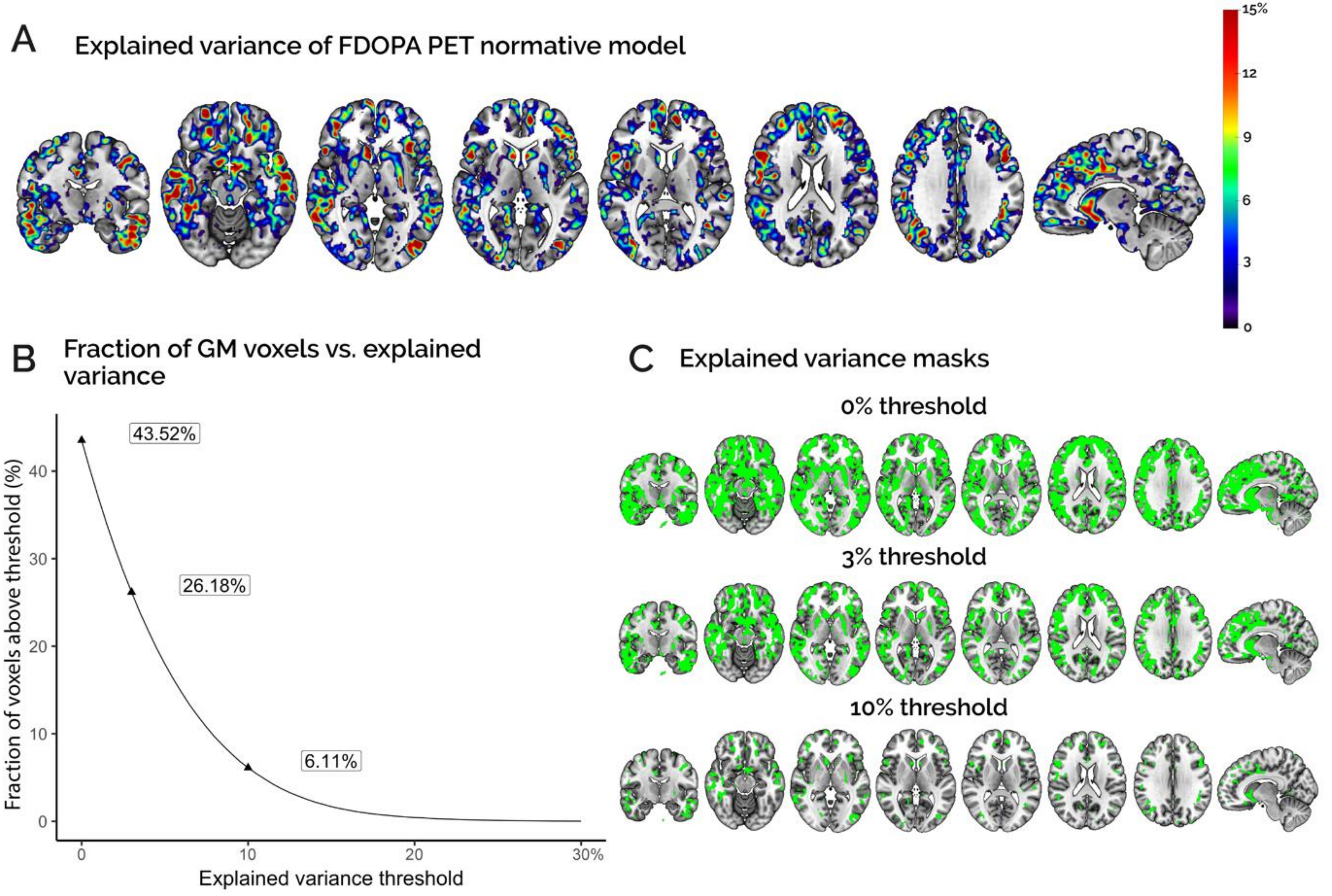

A
Explained variance of FDOPA PET normative model
15%
12
9
6
3
0
B
Fraction of GM voxels vs. explained variance
43.52%
26.18%
6.11%
Fraction of voxels above threshold (%)
0
10
20
30
40
0
10
20
30%
Explained variance threshold
C
Explained variance masks
0% threshold
3% threshold
10% threshold

Supplementary Figure 2. Explained variance of [$^{18}$F]FDOPA PET normative model. (A) Representation of voxel-wise explained variance by the FDOPA normative model in standard MNI coordinates; (B) Percentage of GM voxels as function of different explained variance thresholds: 0% (i.e., wherever the model converges), 3% (i.e., statistical significance threshold given the available sample size, $p<0.05$ uncorrected) and 10% (conservative threshold). (C) Masks of GM voxels defined by FDOPA normative model with different levels of explained variance. These masks exhibit a bilateral distribution and as shown also in panel (B) these masks preserved 43.52%, 26.18% and 6.11% of the original grey matter mask used for the estimation of the normative model.

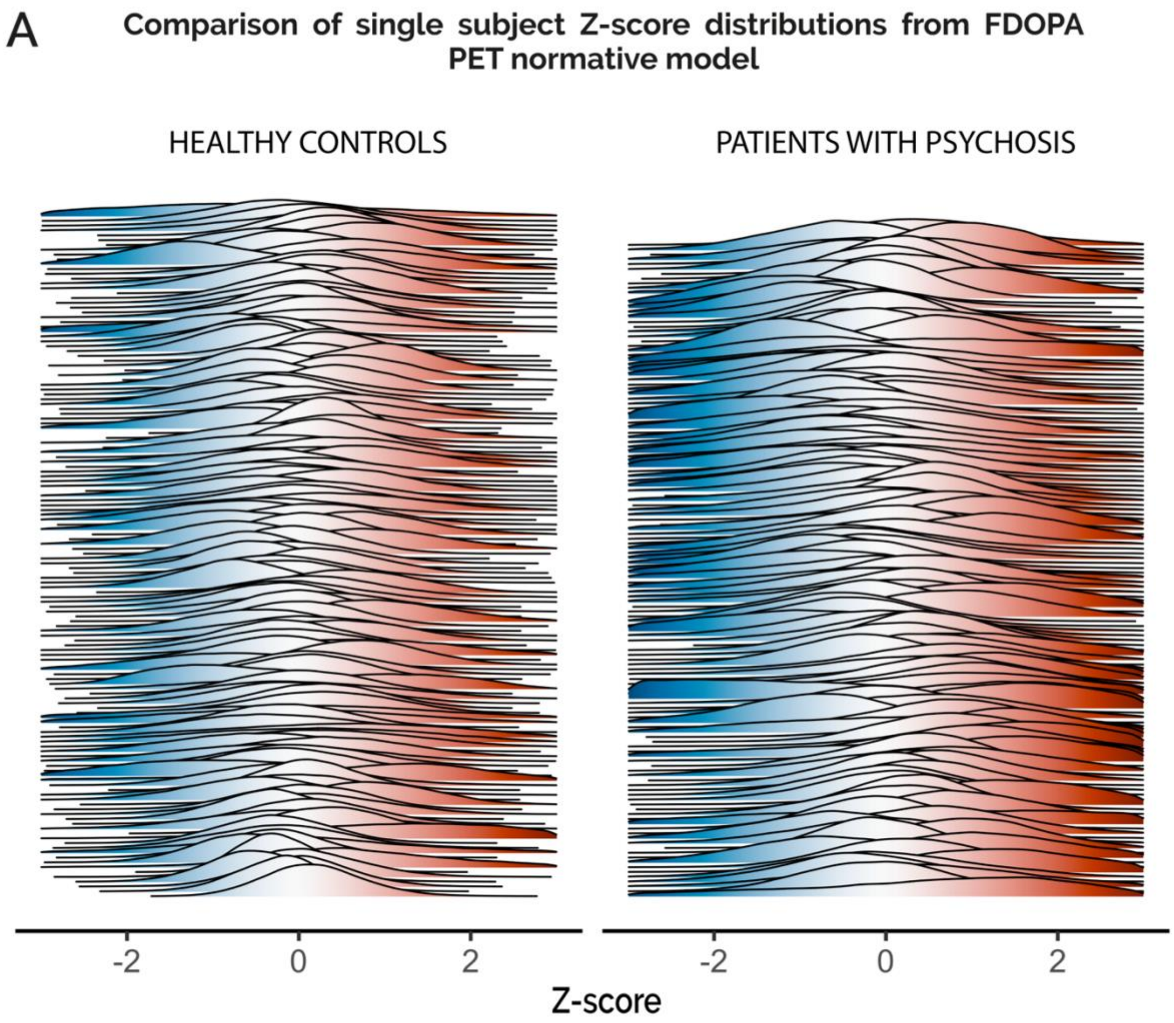


Supplementary Figure 3. Individual level whole brain Z-score distribution of [[18]F]FDOPA PET normative model. (A) Whole brain distribution of Z-scores for each individual of healthy controls (N=109) and patients' cohort (N=136). Z-scores are calculated using k-fold (k=5) cross validation for healthy controls and with the full-model.

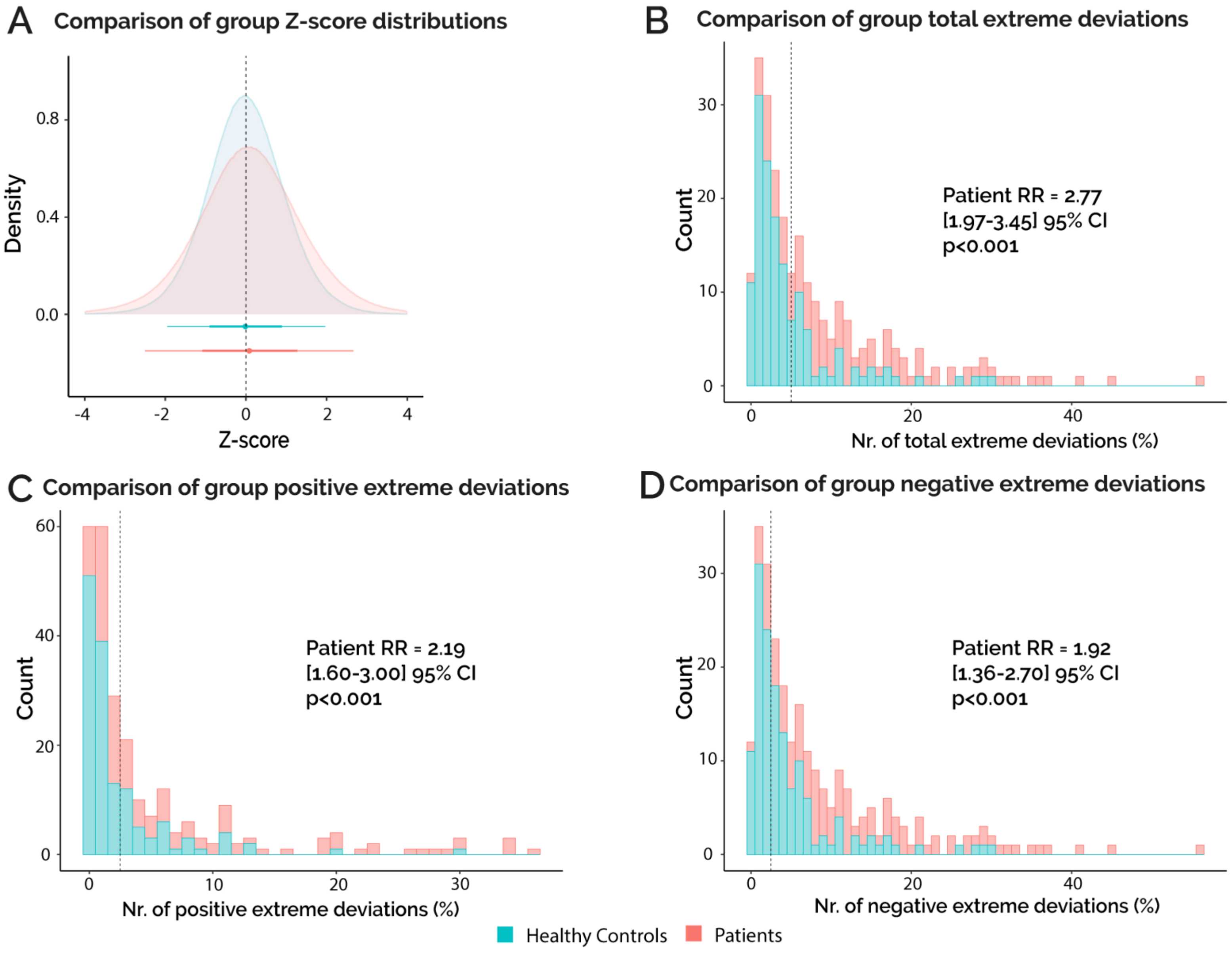
A Comparison of group Z-score distributions
Density
Z-score
B Comparison of group total extreme deviations
Count
Nr. of total extreme deviations (%)
Patient RR = 2.77
[1.97-3.45] 95% CI
p<0.001
C Comparison of group positive extreme deviations
Count
Nr. of positive extreme deviations (%)
Patient RR = 2.19
[1.60-3.00] 95% CI
p<0.001
D Comparison of group negative extreme deviations
Count
Nr. of negative extreme deviations (%)
Patient RR = 1.92
[1.36-2.70] 95% CI
p<0.001
Healthy Controls
Patients

Supplementary Figure 4. Whole brain grouped distributions of Z-scores and extreme deviation scores. (A) Grouped whole brain distribution of Z-scores for healthy controls (N=109) and patients (N=136) cohorts. Z-scores are calculated using k-fold (k=5) cross validation for healthy controls and with the full-model. (B) Percentage of number of whole brain total extreme deviations in healthy controls and patient's cohort. Total extreme deviations are calculated as $|Z|>2$. (C) Percentage of number of whole brain positive extreme deviations in healthy controls and patient cohorts. Positive extreme deviations are calculated as $Z>2$. (D) Percentage of number of whole brain negative extreme deviations in healthy controls and patient cohorts. Negative extreme deviations are calculated as $Z<-2$

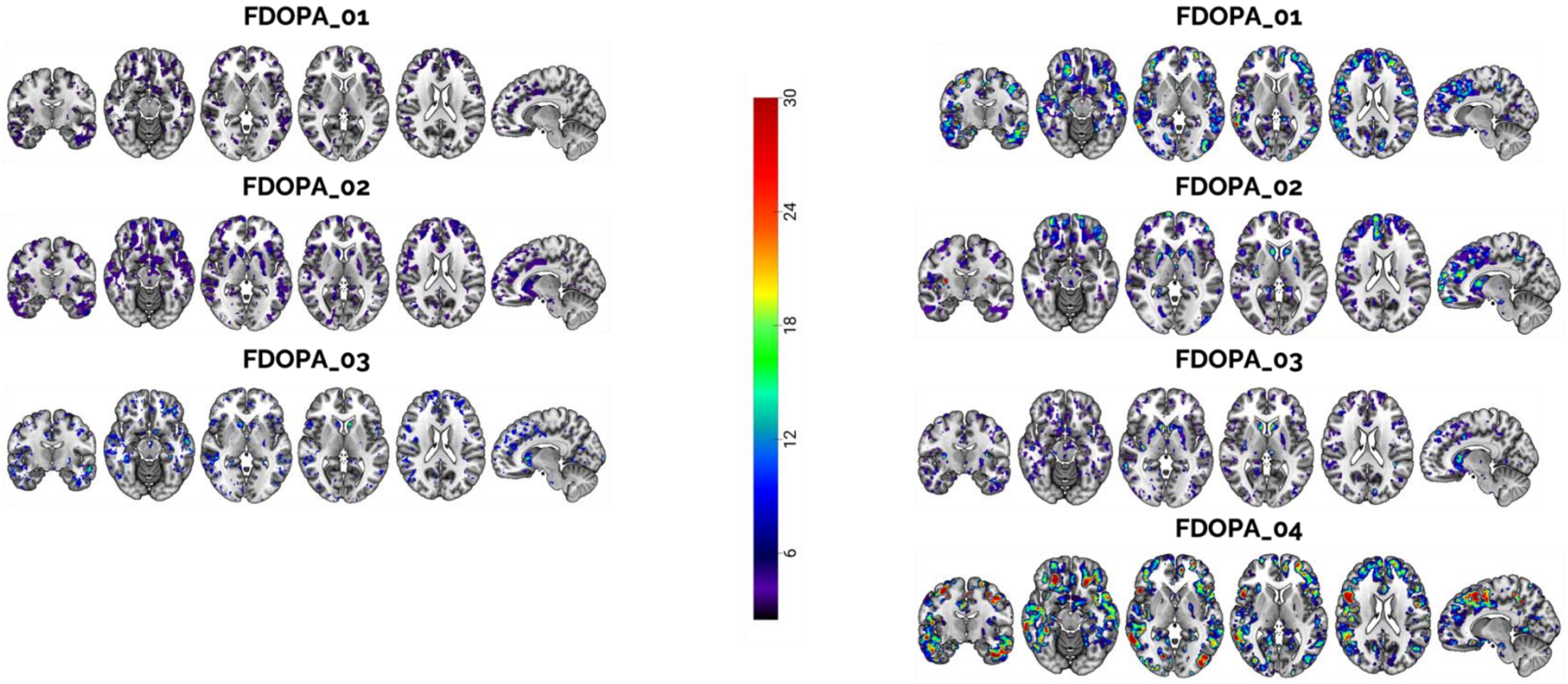


**Supplementary Figure 5: Spatial distribution of extreme positive deviations in study datasets**. Figure shows spatial distribution of extreme positive deviations among HC (left) and patients (right). Maps are thresholded above level of chance (i.e., 2.5%). Study FDOPA_04 has no right column as no matched HC are acquired in the original study. Colorbar indicates the percentage of each cohort which overlaps in the same voxel.

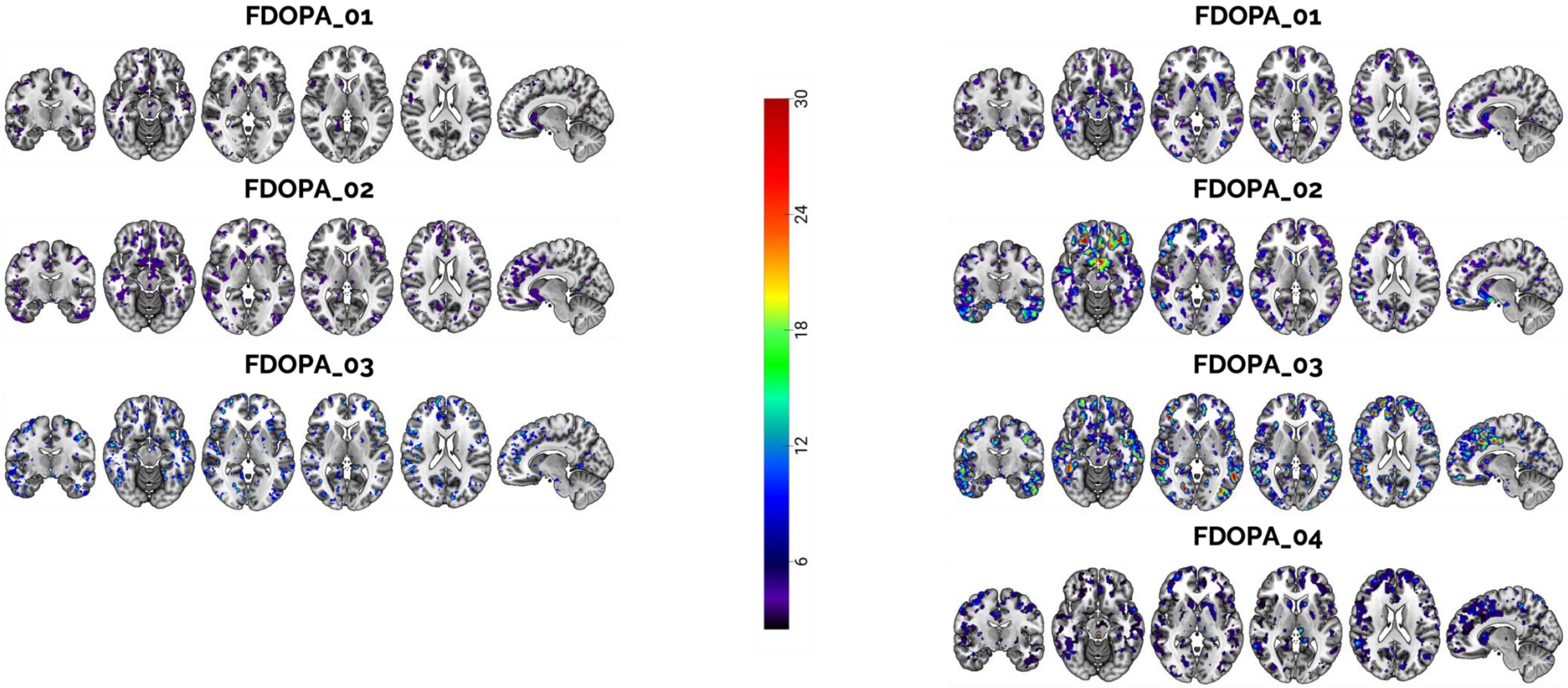


**Supplementary Figure 6: Spatial distribution of extreme negative deviations in study datasets**. Figure shows spatial distribution of extreme positive deviations among HC (left) and patients (right). Maps are thresholded above level of chance (i.e., 2.5%). Study FDOPA_04 has no right column as no matched HC are acquired in the original study. Colorbar indicates the percentage of each cohort which overlaps in the same voxel.

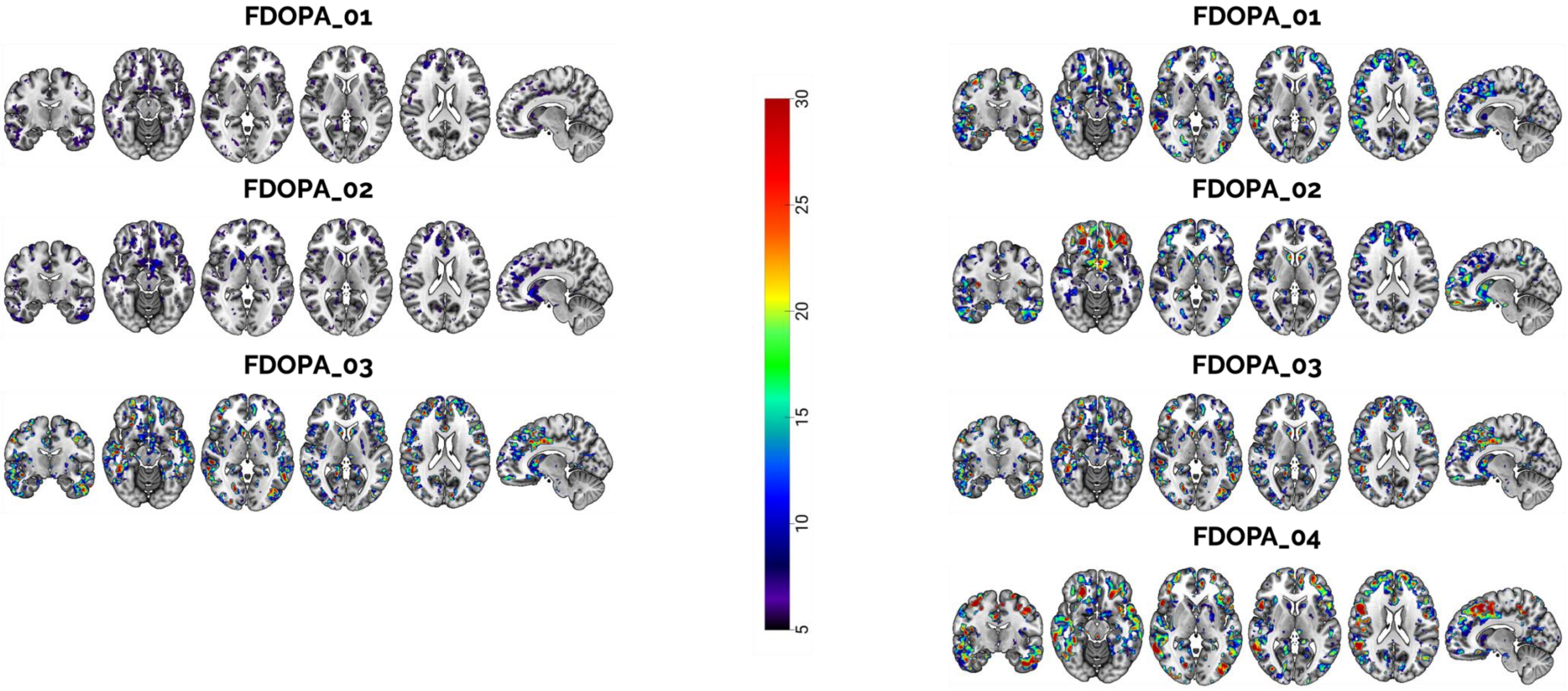


**Supplementary Figure 7: Spatial distribution of extreme total deviations in study datasets**. Figure shows spatial distribution of extreme positive deviations among HC (left) and patients (right). Maps are thresholded above level of chance (i.e., 5%). Study FDOPA_04 has no right column as no matched HC are acquired in the original study. Colorbar indicates the percentage of each cohort which overlaps in the same voxel.

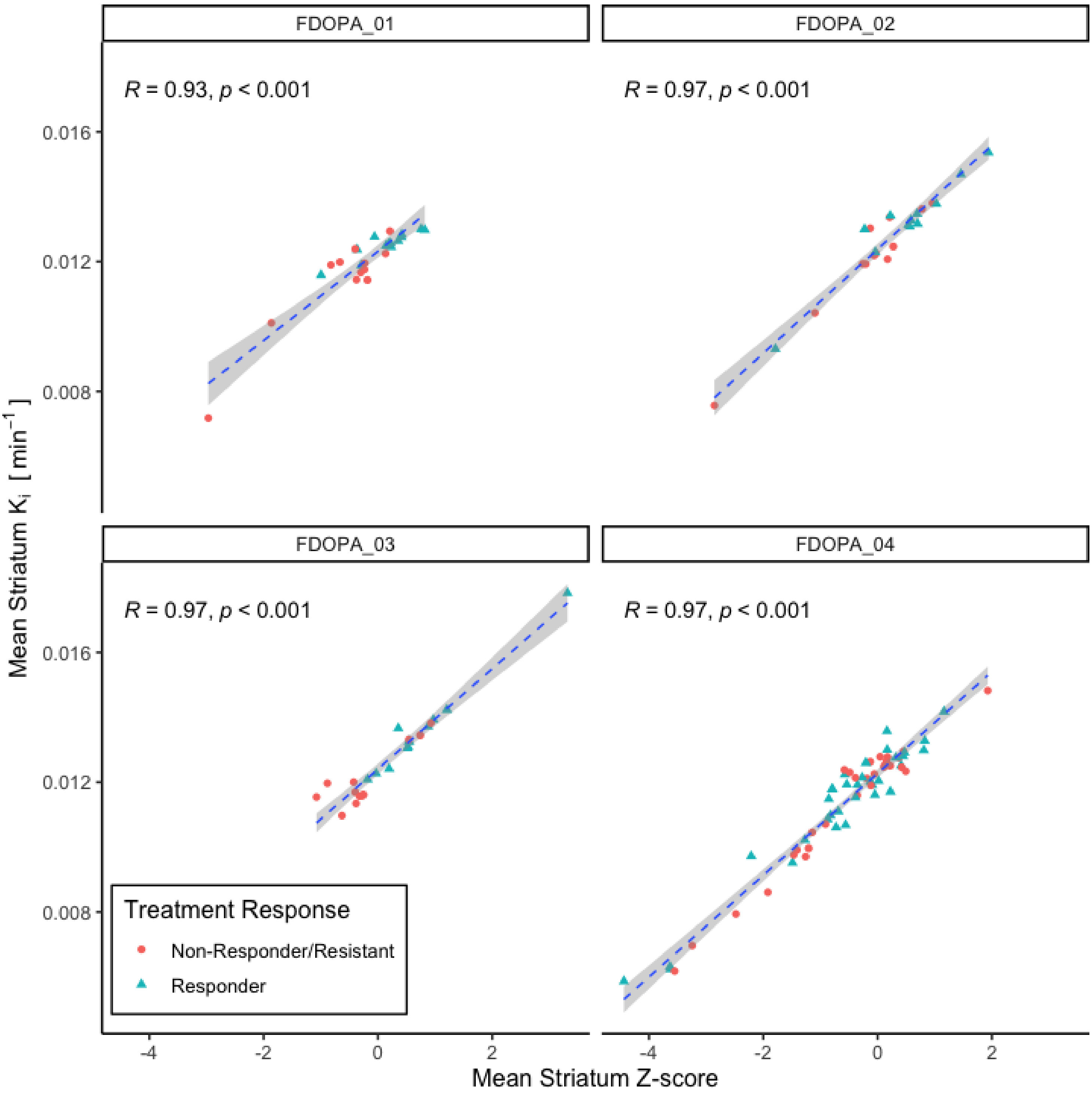


**Supplementary Figure 8: Correlation between striatal $K_i$ and Z-score in patients**. The figure shows the correlation between the average distribution of Z-scores and $K_i^{cer}$ in the striatum (as defined by the Hammersmith atlas) in different cohorts. Dashed lines represent regression lines, while the shaded area shows confidence intervals of the regression. Data markers show patients' response to antipsychotic treatment.

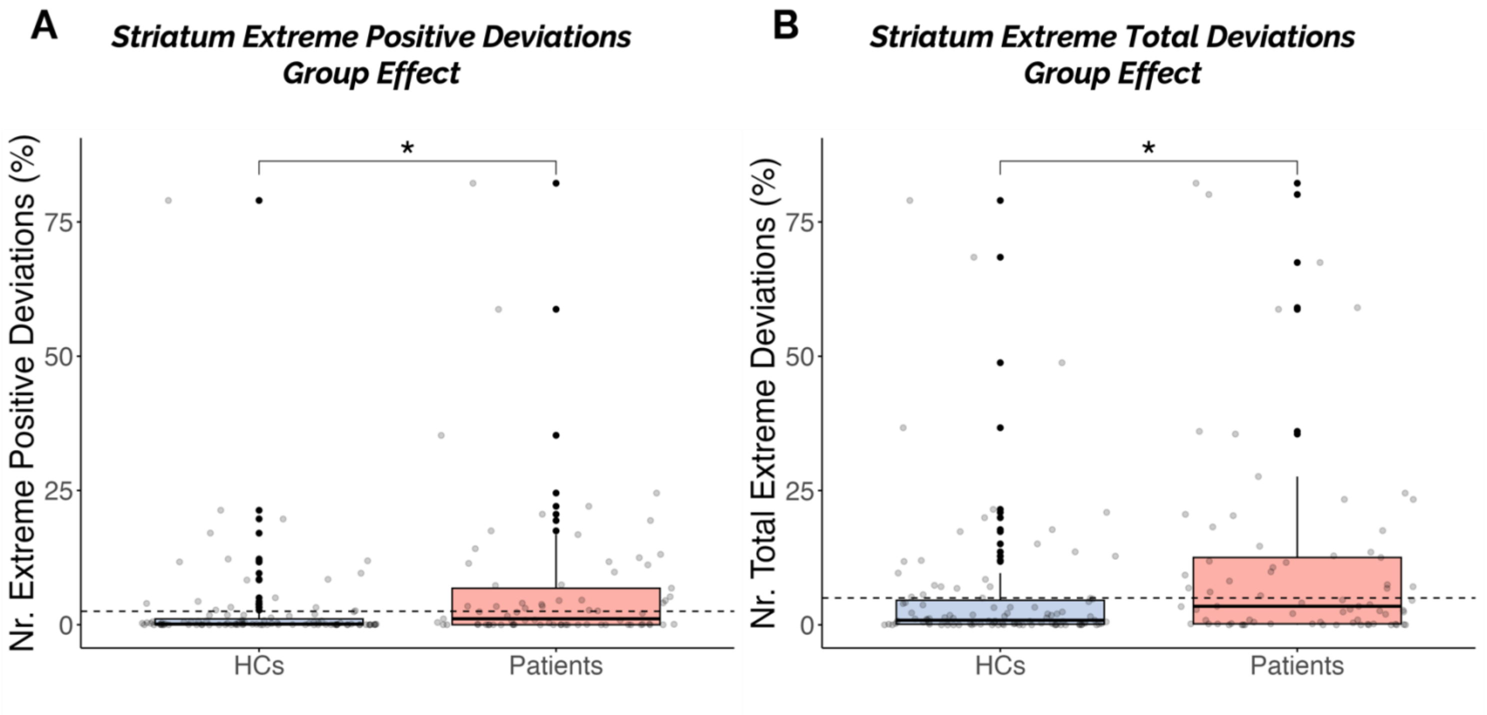


**Supplementary Figure 9: ANOVA analysis of striatal deviation scores**. The figure shows the significant group effects found in the ANOVA analysis of deviation scores in the striatum. (A) Difference of extreme positive deviations between healthy controls and patients (B) Difference of total extreme deviations between groups. Patients are composed of a mixture of first episode (FDOPA_01) and chronic psychosis (FDOPA_02, FDOPA_03), depending on the dataset. Asterisks indicates significance, * indicates $p<0.05$,

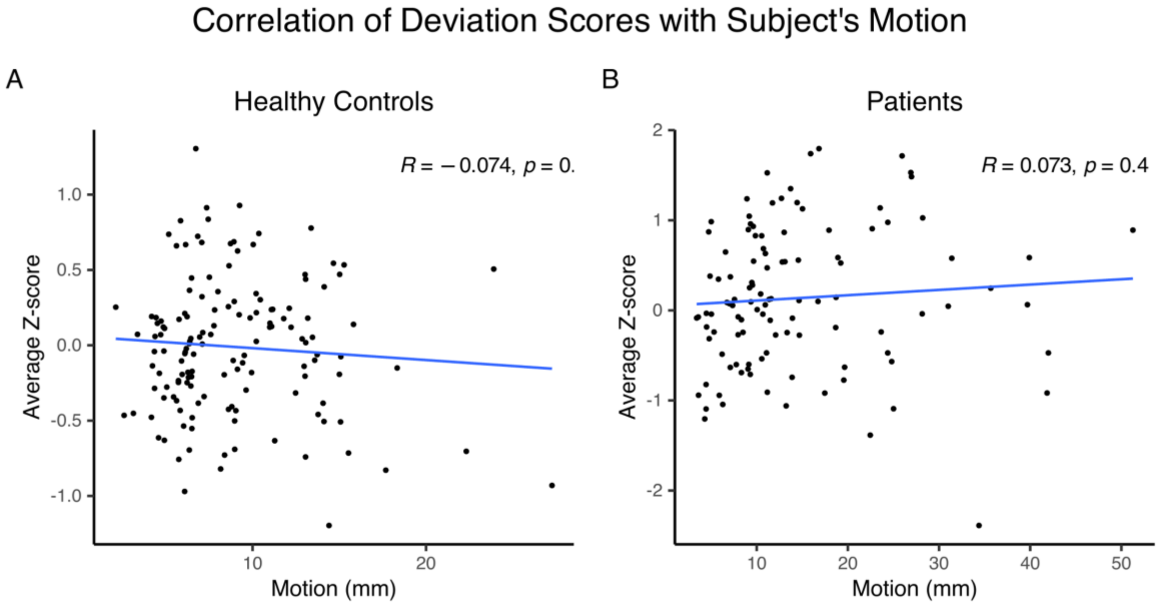


**Supplementary Figure 10: Correlation analysis between Total Patient Motion and Z-score**. The figure shows the Spearman correlation between the subject's total motion (mm) in healthy controls (A) and patients (B). Dots represent individual data points; blue line shows the linear correlation between the two scores.

Supplementary Table 1. ANOVA and Post-hoc t-test Average Z-score [18F]FDOPA – 0% threshold mask. Table shows ANOVA and post-hoc t-test for the interaction between group (HC v. patients) and dataset and the whole brain average Z-score, in the 0% threshold mask.

| VARIABLE | df | F | p |
|---|---|---|---|
| GROUP | 1 | 3.118 | 0.0789 |
| DATASET | **3** | **11.044** | **9.63e-07** |
| INTERACTION | **2** | **5.057** | **0.0072** |

* ANOVA performed in R version 4.2.1 using the anova_test function of the rstatix package

| | | | 95% confidence interval | |
|---|---|---|---|---|
| **DATASETS** | **t** | **p** | **Lower bound** | **Upper bound** |
| FDOPA_01 | **-2.97** | **5.79e-03** | **-0.580** | **-0.109** |
| FDOPA_02 | 0.659 | 0.514 | -0.205 | 0.403 |
| FDOPA_03 | **2.96** | **5.34e-03** | **0.104** | **0.563** |
| FDOPA_04 | n.a. | n.a. | n.a. | n.a. |

*Welch two sample t test between healthy controls and patients performed in R version 4.2.1 using the t_test function in the rstatix package

Supplementary Table 2. ANOVA and Post-hoc t-test Positive Extreme Deviations [18F]FDOPA – 0% threshold mask. Table shows ANOVA and post-hoc t-test for the interaction between group (HC v. patients) and dataset and the number of whole brain extreme positive deviations, i.e., Z>2, in the 0% threshold mask.

| VARIABLE | df | F | p |
|---|---|---|---|
| GROUP | **1** | **30.058** | **1.25e-07** |
| DATASET | **3** | **9.521** | **6.57e-06** |
| INTERACTION | **2** | **3.428** | **0.0344** |

* ANOVA performed in R version 4.2.1 using the anova_test function of the rstatix package

| | | | 95% confidence interval | |
|---|---|---|---|---|
| **DATASETS** | **t** | **p** | **Lower bound** | **Upper bound** |
| FDOPA_01 | **-3.22** | **0.00347** | **-10.5** | **-2.30** |
| FDOPA_02 | -0.543 | 0.59 | -4.98 | 2.86 |
| FDOPA_03 | 0.231 | 0.82 | -2.18 | 2.72 |
| FDOPA_04 | n.a. | n.a. | n.a. | n.a. |

*Welch two sample t test performed in R version 4.2.1 using the t_test function in the rstatix package

Supplementary Table 3. ANOVA and Post-hoc t-test Negative Extreme Deviations [18F]FDOPA – 0% threshold mask. Table shows ANOVA and post-hoc t-tests for group (HC v. patients) and dataset effects and the number of whole brain extreme negative deviations, i.e., Z<-2, in the 0% threshold mask.

| VARIABLE | df | F | p |
|---|---|---|---|
| GROUP | **1** | **16.927** | **5.66e-05** |
| DATASET | **3** | **6.753** | **2.31e-04** |

| INTERACTION | 2 | 2.506 | 0.084129 |
|---|---|---|---|

* ANOVA performed in R version 4.2.1 using the anova_test function of the rstatix package

| | | | 95% confidence interval | |
|---|---|---|---|---|
| **GROUP** | **t** | **p** | **Lower bound** | **Upper bound** |
| HC-PATIENTS | -4.06 | **7.915e-05** | -4.767 | -1.645 |

* Welch two sample t test performed in R version 4.2.1 using the t_test function in the rstatix package

| | | | 95% confidence interval | |
|---|---|---|---|---|
| **DATASETS** | **t** | **p** | **Lower bound** | **Upper bound** |
| FDOPA_03 - FDOPA_01 | 5.355 | **3.51e-06** | 3.693 | 8.164 |
| FDOPA_03 - FDOPA_02 | 3.745 | **3.97e-04** | 2.200 | 7.237 |
| FDOPA_03 - FDOPA_04 | 1.801 | 0.077 | -0.392 | 7.494 |
| FDOPA_01 - FDOPA_02 | -1.620 | 0.110 | -2.698 | 0.279 |
| FDOPA_01 – FDOPA_04 | -1.407 | 0.168 | -5.799 | 1.045 |
| FDOPA_02 – FDOPA_04 | -0.65108 | 0.518 | -4.777 | 2.441 |

*Welch two sample t test performed in R version 4.2.1 using the t_test function in the rstatix package

Supplementary Table 4. ANOVA and Post-hoc t-test Total Extreme Deviations [18F]FDOPA – 0% threshold mask. Table shows ANOVA and post-hoc t-tests for group (HC v. patients) and dataset effects and the number of whole brain total extreme deviations, i.e., |Z|>2, in the 0% threshold mask.

| **VARIABLE** | **df** | **F** | **p** |
|---|---|---|---|
| GROUP | **1** | **53.069** | **7.17e-12** |
| DATASET | **3** | **5.486** | **1.21e-03** |
| INTERACTION | 2 | 0.383 | 0.68215 |

* ANOVA performed in R version 4.2.1 using the anova_test function of the rstatix package

| | | | 95% confidence interval | |
|---|---|---|---|---|
| **GROUP** | **t** | **p** | **Lower bound** | **Upper bound** |
| HC-PATIENTS | -7.276 | **1.298e-11** | -10.815 | -6.198 |

* ANOVA performed in R version 4.2.1 using the anova_test function of the rstatix package

| | | | 95% confidence interval | |
|---|---|---|---|---|
| **DATASETS** | **t** | **p** | **Lower bound** | **Upper bound** |
| FDOPA_03 - FDOPA_01 | **3.938** | **1.92e-04** | **2.686** | **8.201** |
| FDOPA_03 - FDOPA_02 | **2.494** | **0.0146** | **0.843** | **7.479** |
| FDOPA_03 – FDOPA_04 | **-2.430** | **0.0185** | **-10.757** | **-1.030** |
| FDOPA_01 - FDOPA_02 | -0.8840 | 0.379 | -4.165 | 1.600 |
| FDOPA_04 – FDOPA_01 | **4.971** | **1.03e-05** | **6.7430** | **15.932** |
| FDOPA_02 - FDOPA_04 | **-4.080** | **1.42e-04** | **-14.990** | **-5.120** |

*Welch two sample t test performed in R version 4.2.1 using the t_test function in the rstatix package

Supplementary Table 5. ANOVA and Post-hoc t-test Average Z-score [18F]FDOPA – 3% threshold mask. Table shows ANOVA and post-hoc t-test for the interaction between group (HC v. patients) and dataset and the whole brain average Z-score, in the 3% threshold mask.

| VARIABLE | df | F | p |
|---|---|---|---|
| GROUP | 1 | 3.776 | 0.05339 |
| DATASET | **3** | **13.982** | **2.58e-08** |
| INTERACTION | **2** | **5.992** | **0.00297** |

* ANOVA performed in R version 4.2.1 using the anova_test function of the rstatix package

| | | | 95% confidence interval | |
|---|---|---|---|---|
| **DATASETS** | **t** | **p** | **Lower bound** | **Upper bound** |
| FDOPA_01 | **-3.08** | **0.00397** | **-0.644** | **-0.133** |
| FDOPA_02 | 0.779 | 0.44 | -0.196 | 0.442 |
| FDOPA_03 | **3.05** | **0.00485** | **0.129** | **0.655** |
| FDOPA_04 | n.a. | n.a. | n.a. | n.a. |

*Welch two sample t test between healthy controls and patients performed in R version 4.2.1 using the t_test function in the rstatix package.

Supplementary Table 6. ANOVA and Post-hoc t-test Positive Extreme Deviations [18F]FDOPA – 3% threshold mask. Table shows ANOVA and post-hoc t-test for the interaction between group (HC v. patients) and dataset and the number of whole brain extreme positive deviations, i.e., Z>2, in the 3% threshold mask.

| VARIABLE | df | F | p |
|---|---|---|---|
| GROUP | **1** | **30.998** | **8.21e-08** |
| DATASET | **3** | **11.417** | **6.04e-07** |
| INTERACTION | **2** | **3.609** | **0.0289** |

* ANOVA performed in R version 4.2.1 using the anova_test function of the rstatix package

| | | | 95% confidence interval | |
|---|---|---|---|---|
| **DATASETS** | **t** | **p** | **Lower bound** | **Upper bound** |
| FDOPA_01 | **-3.23** | **0.00342** | **-11.6** | **-2.56** |
| FDOPA_02 | -0.480 | 0.634 | -5.09 | 3.13 |
| FDOPA_03 | 0.384 | 0.706 | -2.08 | 2.99 |
| FDOPA_04 | n.a. | n.a. | n.a. | n.a. |

*Welch two sample t test performed in R version 4.2.1 using the t_test function in the rstatix package

Supplementary Table 7. ANOVA and Post-hoc t-test Negative Extreme Deviations [18F]FDOPA – 3% threshold mask. Table shows ANOVA and post-hoc t-test for the interaction between group (HC v. patients) and dataset and the number of whole brain extreme negative deviations, i.e., Z<-2, in the 3% threshold mask.

| VARIABLE | df | F | p |
|---|---|---|---|
| GROUP | **1** | **17.802** | **3.71e-05** |
| DATASET | **3** | **8.676** | **1.93e-05** |
| INTERACTION | **2** | **3.529** | **0.0312** |

* ANOVA performed in R version 4.2.1 using the anova_test function of the rstatix package

| | | | 95% confidence interval | |
|---|---|---|---|---|
| **DATASETS** | **t** | **p** | **Lower bound** | **Upper bound** |
| FDOPA_01 | -0.571 | 0.572 | -1.97 | 1.11 |
| FDOPA_02 | **-2.67** | **0.0112** | **-6.57** | **-0.90** |
| FDOPA_03 | **-3.36** | **2.07e-03** | **-10.9** | **-2.66** |
| FDOPA_04 | n.a. | n.a. | n.a. | n.a. |

*Welch two sample t test between healthy controls and patients performed in R version 4.2.1 using the t_test function in the rstatix package

Supplementary Table 8. ANOVA and Post-hoc t-test Total Extreme Deviations [18F]FDOPA – 3% threshold mask. Table shows ANOVA and post-hoc t-tests for group (HC v. patients) and dataset effects and the number of whole brain total extreme deviations, i.e., |Z|>2, in the 3% threshold mask.

| **VARIABLE** | **df** | **F** | **p** |
|---|---|---|---|
| GROUP | **1** | **56.753** | **1.65e-12** |
| DATASET | **3** | **6.082** | **5.56e-04** |
| INTERACTION | 2 | 0.355 | 0.701476 |

* ANOVA performed in R version 4.2.1 using the anova_test function of the rstatix package

| | | | 95% confidence interval | |
|---|---|---|---|---|
| **GROUP** | **t** | **p** | **Lower bound** | **Upper bound** |
| HC-PATIENTS | **-7.510** | **3.904e-12** | **-11.816** | **-6.895** |

* ANOVA performed in R version 4.2.1 using the anova_test function of the rstatix package

| | | | 95% confidence interval | |
|---|---|---|---|---|
| **DATASETS** | **t** | **p** | **Lower bound** | **Upper bound** |
| FDOPA_03 - FDOPA_01 | **3.989** | **1.641e-04** | **2.958** | **8.880** |
| FDOPA_03 - FDOPA_02 | **2.579** | **0.01167** | **1.044** | **8.084** |
| FDOPA_03 - FDOPA_04 | **-2.543** | **0.01393** | **-11.918** | **-1.408** |
| FDOPA_01 - FDOPA_02 | -0.891 | 0.3752 | -4.376 | 1.665 |
| FDOPA_01 – FDOPA_04 | **5.116** | **6.447e-06** | **7.627** | **17.537** |
| FDOPA_02 – FDOPA_04 | **-4.250** | **8.186e-05** | **-16.518** | **-5.935** |

*Welch two sample t test performed in R version 4.2.1 using the t_test function in the rstatix package

Supplementary Table 9. ANOVA and Post-hoc t-test Average Z-score [18F]FDOPA – 10% threshold mask. Table shows ANOVA and post-hoc t-test for the interaction between group (HC v. patients) and dataset and the whole brain average Z-score, in the 10% threshold mask.

| **VARIABLE** | **df** | **F** | **p** |
|---|---|---|---|
| GROUP | 1 | **5.140** | **0.0244** |
| DATASET | **3** | **21.143** | **6.04e-12** |

| | | | |
|---|---|---|---|
| INTERACTION | **2** | **7.876** | **5.09e-04** |

* ANOVA performed in R version 4.2.1 using the anova_test function of the rstatix package

| | | | 95% confidence interval | |
|---|---|---|---|---|
| **DATASETS** | **t** | **p** | **Lower bound** | **Upper bound** |
| FDOPA_01 | **-3.24** | **2.74e-03** | **-0.741** | **-0.169** |
| FDOPA_02 | 1.13 | 0.264 | -0.149 | 0.531 |
| FDOPA_03 | **-3.33** | **2.42e-03** | **-13.8** | **-3.29** |
| FDOPA_04 | n.a. | n.a. | n.a. | n.a. |

*Welch two sample t test performed in R version 4.2.1 using the t_test function in the rstatix package

Supplementary Table 10. ANOVA and Post-hoc t-test Positive Extreme Deviations [18F]FDOPA – 10% threshold mask. Table shows ANOVA and post-hoc t-test for the interaction between group (HC v. patients) and dataset and the number of whole brain extreme positive deviations, i.e., Z>2, in the 10% threshold mask.

| **VARIABLE** | **df** | **F** | **p** |
|---|---|---|---|
| GROUP | **1** | **33.355** | **2.88e-08** |
| DATASET | **3** | **15.242** | **5.65e-09** |
| INTERACTION | **2** | **3.622** | **0.0285** |

* ANOVA performed in R version 4.2.1 using the anova_test function of the rstatix package

| | | | 95% confidence interval | |
|---|---|---|---|---|
| **DATASETS** | **t** | **p** | **Lower bound** | **Upper bound** |
| FDOPA_01 | **-3.26** | **0.00321** | **-13.7** | **-3.08** |
| FDOPA_02 | -0.384 | 0.703 | -5.26 | 3.58 |
| FDOPA_03 | 0.545 | 0.594 | -2.16 | 3.64 |
| FDOPA_04 | n.a. | n.a. | n.a. | n.a. |

*Welch two sample t test performed in R version 4.2.1 using the t_test function in the rstatix package

Supplementary Table 11. ANOVA and Post-hoc t-test Negative Extreme Deviations [18F]FDOPA – 10% threshold mask. Table shows ANOVA and post-hoc t-test for the interaction between group (HC v. patients) and dataset and the number of whole brain extreme negative deviations, i.e., Z<-2, in the 3% threshold mask.

| **VARIABLE** | **df** | **F** | **p** |
|---|---|---|---|
| GROUP | **1** | **23.011** | **3.13e-06** |
| DATASET | **3** | **14.816** | **9.42e-09** |
| INTERACTION | **2** | **5.981** | **0.003** |

* ANOVA performed in R version 4.2.1 using the anova_test function of the rstatix package

| | | | 95% confidence interval | |
|---|---|---|---|---|
| **DATASETS** | **t** | **p** | **Lower bound** | **Upper bound** |
| FDOPA_01 | -0.476 | 0.637 | -1.98 | 1.23 |

| | | | | |
|---|---|---|---|---|
| FDOPA_03 | -3.24 | 0.00279 | -8.54 | -1.95 |
| FDOPA_04 | -3.33 | 0.00242 | -13.8 | -3.29 |
| FDOPA_02 | n.a. | n.a. | n.a. | n.a. |

*Welch two sample t test performed in R version 4.2.1 using the t_test function in the rstatix package

Supplementary Table 12. ANOVA and Post-hoc t-test Total Extreme Deviations [18F]FDOPA – 10% threshold mask. Table shows ANOVA and post-hoc t-tests for group (HC v. patients) and dataset effects and the number of whole brain total extreme deviations, i.e., |Z|>2, in the 10% threshold mask.

| **VARIABLE** | **df** | **F** | **p** |
|---|---|---|---|
| GROUP | **1** | **65.110** | **6.33e-14** |
| DATASET | **3** | **7.299** | **1.14e-04** |
| INTERACTION | **2** | 0.248 | 0.780 |

* ANOVA performed in R version 4.2.1 using the anova_test function of the rstatix package

| | | | 95% confidence interval | |
|---|---|---|---|---|
| **GROUP** | **t** | **p** | **Lower bound** | **Upper bound** |
| HC-PATIENTS | -8.01 | **2.98e-13** | -14.29 | -8.63 |

* ANOVA performed in R version 4.2.1 using the anova_test function of the rstatix package

| | | | 95% confidence interval | |
|---|---|---|---|---|
| **DATASETS** | **t** | **p** | **Lower bound** | **Upper bound** |
| FDOPA_03 - FDOPA_01 | **4.22** | **7.729e-05** | **3.79** | **10.61** |
| FDOPA_03 - FDOPA_02 | **2.72** | **7.961e-04** | **1.47** | **9.45** |
| FDOPA_03 - FDOPA_04 | **-2.71** | **9.136e-03** | **-14.70** | **-2.19** |
| FDOPA_01 - FDOPA_02 | -1.037 | 0.3025 | -5.08 | 1.59 |
| FDOPA_01 – FDOPA_04 | **5.34** | **3.113e-06** | **9.76** | **21.54** |
| FDOPA_02 – FDOPA_04 | **-4.48** | **4.063e-05** | **-20.13** | **-7.68** |

*Welch two sample t test performed in R version 4.2.1 using the t_test function in the rstatix package
